# Retention in care and antiretroviral therapy adherence among Medicaid beneficiaries with HIV, 2001-2015

**DOI:** 10.1101/2024.05.13.24307278

**Authors:** Jacqueline E. Rudolph, Keri L. Calkins, Xueer Zhang, Yiyi Zhou, Xiaoqiang Xu, Eryka L. Wentz, Corinne E. Joshu, Bryan Lau

## Abstract

Disparities in HIV care by socioeconomic status, place of residence, and race/ethnicity prevent progress toward epidemic control. No study has comprehensively characterized the HIV care cascade among people with HIV enrolled in Medicaid – an insurance source for low-income individuals in the US. We analyzed data from 246,127 people with HIV enrolled in Medicaid 2001-2015, aged 18-64, living in 14 US states. We estimated monthly prevalence of four steps of the care cascade: retained in care/adherent to ART; retained/not adherent; not retained/adherent; not retained/not adherent. Beneficiaries were retained in care if they had an outpatient care encounter every six months. Adherence was based on medication possession ratio. We estimated prevalence using a non-parametric multi-state approach, accounting for death as a competing event and for Medicaid disenrollment using inverse probability of censoring weights. Across 2001-2015, the proportion of beneficiaries with HIV who were retained/ART adherent increased, overall and in all subgroups. By 2015, approximately half of beneficiaries were retained in care, and 42% of beneficiaries were ART adherent. We saw meaningful differences by race/ethnicity and region. Our work highlights an important disparity in the HIV care cascade by insurance status during this time period.

## INTRODUCTION

The HIV care cascade is one metric to track progress in controlling the HIV epidemic in a target population. The cascade quantifies the proportion of people with HIV (PWH) in a population who are diagnosed with HIV, linked to care, receiving HIV medical care, retained in care, and virally suppressed (HIV.gov, 2022). Ideally, all PWH would fall into the last category, both for the long-term health of current PWH and to prevent HIV transmission (Bavinton et al., 2018; Cohen et al., 2011; Rodger et al., 2016). In 2019, the Centers for Disease Control (CDC) estimated that 86.7% of PWH in the United States (US) were diagnosed, 65.9% had received HIV care, 50.1% were retained in care, and 56.8% had achieved viral suppression (Centers for Disease Control and Prevention, 2021). While this proportion is a meaningful improvement from the estimated 28% virally suppressed in 2010, the US remains far from the goal set by the “Ending the HIV Epidemic in the US” initiative of achieving 75% suppression by 2025 and 90% suppression by 2030 (Centers for Disease Control and Prevention (CDC), 2011; Office of Infectious Disease and HIV/AIDS Policy, HHS, 2023).

One barrier to achieving these goals are lingering disparities in linkage to and retention in care by race/ethnicity, HIV transmission category, place of residence, and socioeconomic status (Althoff et al., 2014; Centers for Disease Control and Prevention, 2021; Hanna et al., 2013; Lillie-Blanton et al., 2010; Muthulingam et al., 2013; Rebeiro et al., 2017). According to the CDC’s 2019 report, although white PWH in the US have achieved 63.7% viral suppression, suppression among Black (52.6%) and Hispanic PWH (54.0%) lags behind (Centers for Disease Control and Prevention, 2021). Additionally, people who acquired HIV through injection drug use are less likely to be linked to care than people who fall into other transmission categories, and PWH who live in rural areas are also more likely to have AIDS at time of HIV diagnosis (25.2%) than PWH living in metropolitan areas (19.8%) (Centers for Disease Control and Prevention, 2021).

Medicaid is a joint federal and state-administered public insurance for people in the US who are low income or have disabilities. Medicaid is the largest single source of insurance for PWH in the US, with covering care for approximately 40% of PWH (Roberts et al., 2023). While the care cascade is well characterized among all PWH in the US and among those linked to HIV care clinics, less is known about the state of the cascade among PWH on Medicaid (Byrd et al., 2015; Health Resources and Services Administration, 2016). Some prior literature has reported that PWH on Medicaid have lower quality of care, are less likely to be virally suppressed or retained in care, and are more likely to discontinue antiretroviral therapy (ART) than PWH on Medicare or private insurance (Horberg et al., 2020; Komandt et al., 2023; Ludema et al., 2016; Mugavero et al., 2007; Robison et al., 2008; Yehia et al., 2015). In light of this gap, we characterized the care cascade among PWH enrolled in Medicaid between 2001-2015 in 14 US states, overall and by sex, race/ethnicity, and region. All beneficiaries identified as having HIV through claims records are aware of their diagnosis and linked to care; thus, we focused on the later steps of the care cascade, namely retention in care and adherence to ART.

## MATERIALS AND METHODS

### Study Sample

We used 2001-2015 Medicaid Analytic eXtract and Transformed Medicaid Statistical Information System Analytical Files data from the Centers for Medicare and Medicaid Services for beneficiaries enrolled in 14 US states: Alabama (AL), California (CA), Colorado (CO), Florida (FL), Georgia (GA), Illinois (IL), Maryland (MD), Massachusetts (MA), New York (NY), North Carolina (NC), Ohio (OH), Pennsylvania (PA), Texas (TX), and Washington (WA). These data have been described elsewhere (Rudolph et al., 2023). Here, we analyzed data from each Medicaid beneficiary’s first period of continuous enrollment (without dual enrollment in Medicare or private insurance) that exceeded 6 months in length. We included beneficiaries who were aged 18-64 years, had full Medicaid benefits (i.e., all healthcare covered by Medicaid), and had evidence of an HIV diagnosis during this period of enrollment. Baseline for the analysis was the first of the month following a beneficiary’s HIV diagnosis. HIV diagnoses were based on the presence of one inpatient claim or two outpatient claims (required to be within two years) with an HIV-related diagnosis code (see Table S1 for codes) (Chronic Conditions Data Warehouse, n.d.; Fultz et al., 2006; Keller et al., 2014). The date of the first claim was used as the date of diagnosis.

The Johns Hopkins Bloomberg School of Public Health Institutional Review Board (FWA #00000287) determined that this secondary analysis of Medicaid claims data was exempt (IRB no. 11617). No informed consent was required.

### Measures

To be considered retained in care, a beneficiary had to have at least one outpatient claim for an office visit, viral load measurement, or CD4 cell count every 6 months. In each month of follow-up, we determined adherence to ART by using the prescription fill date and days supply to compute the medication possession ratio (MPR), or the proportion of days in the month with ART supply. In our main analysis, a beneficiary was considered adherent if their MPR was 80% or higher. We considered alternate definitions of ART adherence, including if a beneficiary had any evidence of ART (MPR>0), a MPR of at least 70%, or at least 90%.

We included in our analysis covariates we hypothesized would be related with length of Medicaid enrollment and with engagement in HIV care (Anderson et al., 2022; Garfield et al., n.d.; Ku & Platt, 2022). These included age, sex (female, male), race/ethnicity (non-Hispanic Black, non-Hispanic white, Hispanic, Other), US Census region (South: AL, FL, GA, MD, NC, TX; Northeast: MA, NY, PA; Midwest: IL, OH; West: CA, CO, WA), calendar period (2001-2005, 2006-2010, 2011-2015), and number of Charlson comorbidities (0, 1, 2+) (US Census Bureau, 2021). The number of comorbidities was treated as a marker of multimorbidity burden and healthcare needs. The Charlson comorbidities included myocardial infarction, congestive heart failure, peripheral vascular disease, cerebrovascular disease, dementia, chronic pulmonary disease, rheumatic disease, peptic ulcer disease, mild liver disease, diabetes without chronic complication, diabetes with chronic complication, hemiplegia or paraplegia, renal disease, malignancy, and metastatic solid tumor (Charlson et al., 1987; Quan et al., 2011).

### Statistical analysis

From baseline, follow-up was divided into monthly intervals. Beneficiaries were censored when they reached 65 years of age (due to eligibility for Medicare), had evidence of dual enrollment in Medicare or private insurance, or lost Medicaid coverage (Centers for Medicare and Medicaid Services, n.d.). Due to concerns of prescription data quality based on a preliminary assessment of claims completeness, beneficiaries from NC, NY, and MD were administratively censored at the end of 2012, 2013, and 2014, respectively. Otherwise, beneficiaries were administratively censored on September 30, 2015 – the last date for ICD-9 coding.

In each month of follow-up, we categorized a beneficiary as being in one of four levels of the care cascade: ART adherent and retained in care, ART adherent and not retained in care, not ART adherent and retained in care, and not ART adherent and not retained in care. We additionally assessed whether a beneficiary died in a given month of follow-up. We used a non-parametric multi-state approach (specifically, the Aalen-Johansen estimator) to estimate the prevalence of each care cascade category and incidence of death as a competing event, using calendar time as the time scale. This approach has previously been used to investigate the HIV care cascade in other settings (Lesko et al., 2016). To control for possible non-random censoring, we weighted the multi-state model using inverse probability of censoring weights (Buchanan et al., 2014; Cole & Hernan, 2008; Cole & Hernán, 2004). These weights included all covariates mentioned above. We additionally carried out analyses stratified by sex, select race/ethnicity categories (non-Hispanic Black, non-Hispanic white, and Hispanic), and US Census region.

For each year, we report the prevalence of each care cascade category and the cumulative incidence of death on December 31, as well as the increase in the cumulative incidence of death since the prior year. For comparability with prior literature, we also report the prevalence of each care cascade category if we do not consider the cumulative incidence of death incidence, i.e., the estimated prevalence divided by one minus the incidence of mortality. We focus on the results from 2002 onwards, given that no one could be classified as not retained in care in the first 6 months after their HIV diagnosis (which most impacts results for 2001).

We carried out two sensitivity analyses. First, we ran an analysis excluding the state of CA, due to potential underreporting of mortality noted in our preliminary data quality assessment. Second, we report the findings when NY, NC, and MD had the same administrative censoring date as all other states. All analyses were carried out using R version 4.0.5 (R Foundation, Vienna, Austria). Code for this analysis is available on GitHub [BLINDED].

## RESULTS

Our analysis included 246,127 Medicaid beneficiaries with HIV, with a median follow-up of 18 months (IQR: 7, 52). At baseline, the median age was 42.6 (IQR: 34.7, 50.1), and 40.4% of beneficiaries were female (Table 1). The predominant race/ethnicity was non-Hispanic Black (49.8%), followed by non-Hispanic white (23.9%), other races (16.8%), and Hispanic (9.5%). NY was the largest contributor of beneficiaries with HIV (32.3%); other large states included California (16.5%) and Florida (12.6%). Comorbidity burden at baseline was high given the age of our sample, with 23.7% of beneficiaries having one comorbidity and 16.8% having two or more. Looking across follow-up (Table 1), we saw a higher proportion of female beneficiaries contributed time in the retained/not ART adherent state (49.2% of person-months) and a lower proportion contributed time to the not retained/ART adherent state (38.7%). Non-Hispanic Black beneficiaries were more likely to contribute time to the not retained/not ART adherent state (57.8%). Beneficiaries in the retained/ART adherent state were the least likely to be comorbidity-free (28.3%).

**Table 1.** Characteristics of Medicaid beneficiaries with HIV, overall at baseline and across months contributed to states in the care cascade.

| Variable |  | Overall<br>(n=246,127) | Retained<br>ART adherent<br>(m=3,346,396) | Retained<br>Not ART adherent<br>(m=3,386,885) | Not retained<br>ART adherent<br>(m=634,065) | Not retained<br>Not ART adherent<br>(m=1,682,148) |
| --- | --- | --- | --- | --- | --- | --- |
| Age, median (IQR) |  | 42.6 (34.7, 50.1) | 42.9 (36.9, 48.7) | 41.6 (34.8, 48.1) | 42.3 (36.5, 48.1) | 40.9 (34.0, 47.2) |
| Sex |  |  |  |  |  |  |
|  | Female | 110,506 (40.4) | 1,423,440 (40.3) | 1,924,805 (49.2) | 245,355 (38.7) | 805,757 (46.3) |
|  | Male | 152,316 (61.9) | 1,997,440 (59.7) | 1,752,572 (51.7) | 388,108 (61.2) | 895,165 (53.2) |
| Race/ethnicity |  |  |  |  |  |  |
|  | NH Black | 122,487 (49.8) | 1,576,754 (47.1) | 1,812,316 (53.5) | 341,908 (53.9) | 972,545 (57.8) |
|  | NH white | 58,839 (23.9) | 807,813 (24.1) | 725,641 (21.4) | 108,917 (17.2) | 299,577 (17.8) |
|  | Hispanic | 23,358 (9.5) | 333,533 (10.0) | 294,143 (8.7) | 58,334 (9.2) | 136,801 (8.1) |
|  | Other | 41,443 (16.8) | 628,296 (18.8) | 554,785 (16.4) | 124,906 (19.7) | 273,225 (16.2) |
| US State |  |  |  |  |  |  |
| South | AL | 2899 (1.2) | 42,000 (1.3) | 55,318 (1.6) | 2271 (0.4) | 17,169 (1.0) |
|  | FL | 30,980 (12.6) | 478,890 (14.3) | 418,383 (12.4) | 35,700 (5.6) | 177,996 (10.6) |
|  | GA | 11,442 (4.6) | 139,333 (4.2) | 170,894 (5.0) | 7621 (1.2) | 52,799 (3.1) |
|  | MD | 11,392 (4.6) | 84,926 (2.5) | 122,671 (3.6) | 54,553 (8.6) | 156,903 (9.3) |
|  | NC | 8473 (3.4) | 94,679 (2.8) | 128,504 (3.8) | 4998 (0.8) | 30,180 (1.8) |
|  | TX | 14,339 (5.8) | 170,341 (5.1) | 231,892 (6.8) | 20,884 (3.3) | 74,244 (4.4) |
| Northeast | MA | 9800 (4.0) | 122,656 (3.7) | 126,811 (3.7) | 7027 (1.1) | 49,248 (2.9) |
|  | NY | 79,609 (32.3) | 1,303,296 (38.9) | 1,257,270 (37.1) | 318,611 (50.2) | 687,656 (40.9) |
|  | PA | 8670 (3.5) | 75,109 (2.2) | 68,157 (2.0) | 5638 (0.9) | 68,747 (4.1) |
| Midwest | IL | 15,259 (6.2) | 182,321 (5.4) | 214,062 (6.3) | 54,947 (8.7) | 124,846 (7.4) |
|  | OH | 6822 (2.8) | 23,518 (0.7) | 106,644 (3.1) | 1472 (0.2) | 41,641 (2.5) |
| West | CA | 40,548 (16.5) | 583,969 (17.5) | 447,074 (13.2) | 115,911 (18.3) | 186,863 (11.1) |
|  | CO | 1180 (0.5) | 11,858 (0.4) | 10,739 (0.3) | 2763 (0.4) | 9531 (0.6) |
|  | WA | 4714 (1.9) | 33,500 (1.0) | 28,466 (0.8) | 1669 (0.3) | 4325 (0.3) |
| Enrollment period |  |  |  |  |  |  |
|  | 2001-2005 | 123,058 (50.0) | 2,347,478 (70.1) | 2,502,523 (73.9) | 499,077 (78.7) | 1,314,829 (78.2) |
|  | 2006-2010 | 50,435 (20.5) | 572,590 (17.1) | 544,235 (16.1) | 89,301 (14.1) | 258,594 (15.4) |
|  | 2011-2015 | 72,634 (29.5) | 426,328 (12.7) | 340,127 (10.0) | 45,687 (7.2) | 108,725 (6.5) |
| No. comorbidities |  |  |  |  |  |  |
|  | 0 | 146,507 (59.5) | 947,598 (28.3) | 1,132,660 (33.4) | 209,536 (33.0) | 594,262 (35.3) |
|  | 1 | 58,347 (23.7) | 884,858 (26.4) | 922,716 (27.2) | 176,346 (27.8) | 475,471 (28.3) |
|  | 2+ | 41,273 (16.8) | 1,513,940 (45.2) | 1,331,509 (39.3) | 248,183 (39.1) | 612,415 (36.4) |

Abbreviations: ART, antiretroviral therapy; IQR, interquartile range; m, number of months; n, number of beneficiaries; NH, non-Hispanic; No., number of

Overall, 22.1% of beneficiaries with HIV who were still alive were retained/ART adherent in 2002; the proportion retained/ART adherent increased steadily across the study period, reaching a peak of 35.7% in 2014 (Table 2, Figure 1A). When not incorporating the incidence of death, those proportions dropped to 23.1% in 2002 and 48.1% in 2014 (Figure 1B). The proportion not retained in care/ART adherent did not meaningfully change across follow-up. Combining the categories, we estimated that at most 42.1% (57.2% without death) of Medicaid beneficiaries were ART adherent at any point in follow-up. In contrast, the proportion retained in care/not ART adherent decreased from 43.2% (45.1% without death) in 2002 to 14.9% (20.2% without death) in 2015. The proportion not retained in care/not ART adherent decreased from 24.8% (25.9% without death) in 2002 to a minimum of 10.5% (13.9% without death) in 2013, with a slight increase in prevalence 2014-2015. We saw that 17,155 beneficiaries died while enrolled in Medicaid. The annual increase in the cumulative incidence of death decreased over time, from a 2.8% increase between the end of 2001 and the end of 2002 to of a 0.6% increase between 2014-2015.

**Figure 1.**
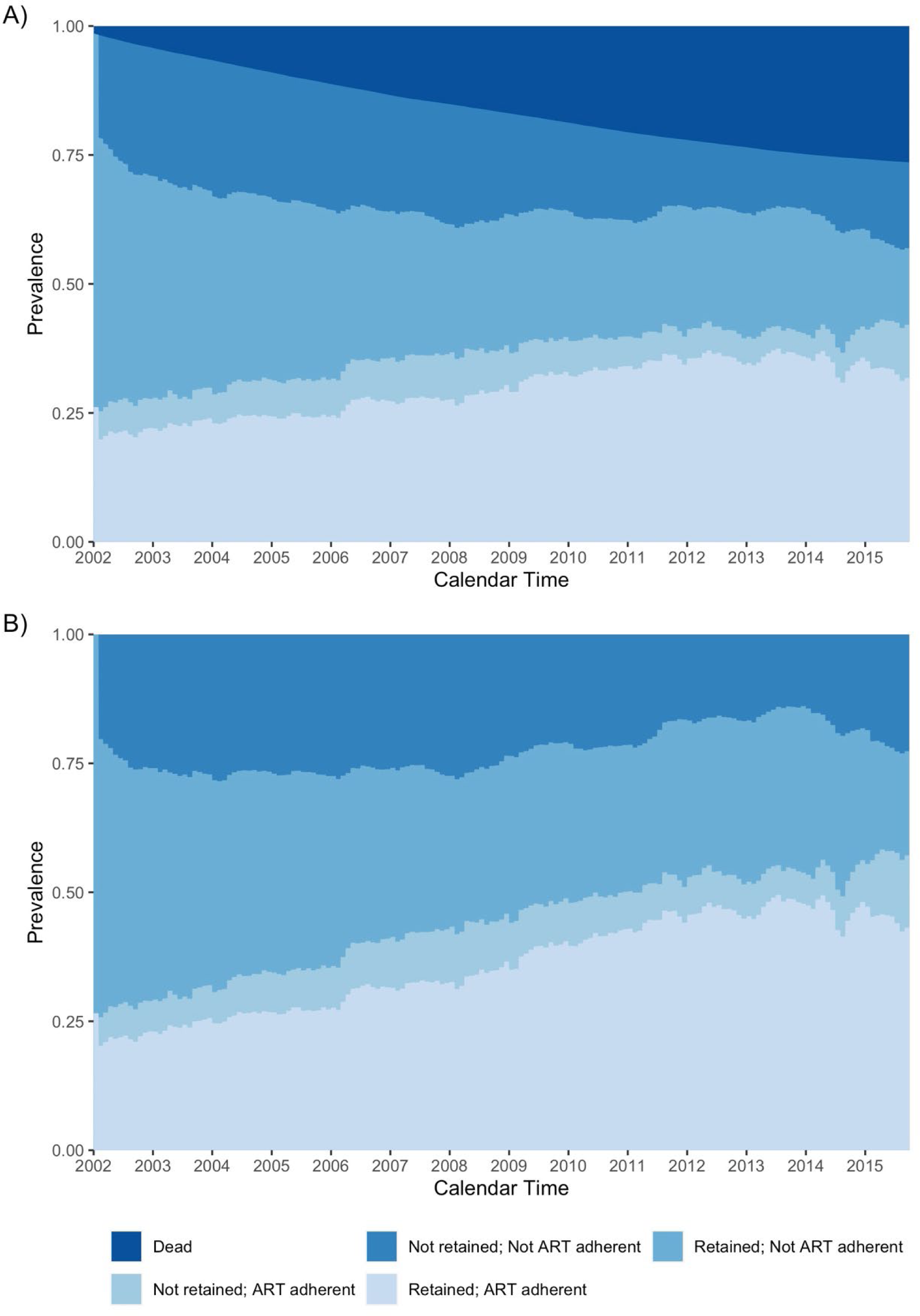
Weighted prevalence of HIV care cascade levels among Medicaid beneficiaries (A) including and (B) not including incidence of death, using an adherence level of 80%.

**Table 2.**
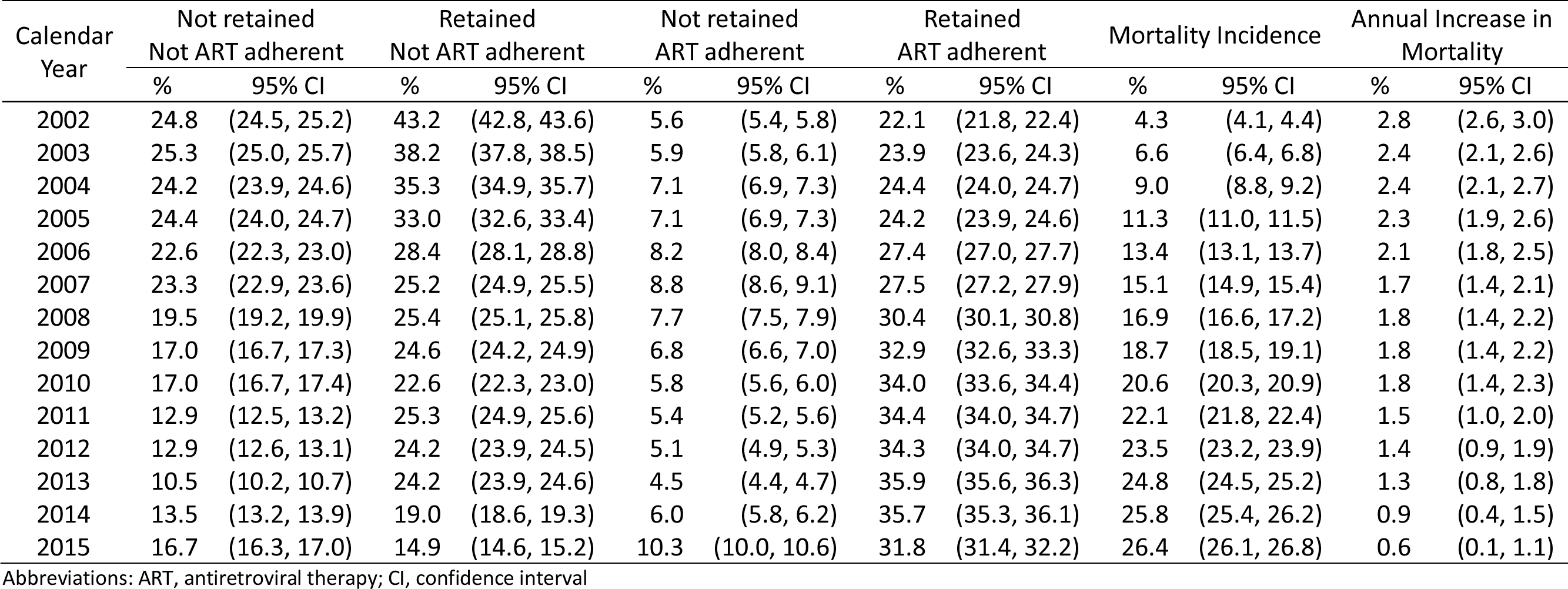
Weighted prevalence of each level of the HIV care cascade and cumulative incidence of mortality by end of the calendar year and the annual increase in the cumulative incidence of mortality, using an adherence level of 80%.

Female beneficiaries were less likely to be ART adherent than male beneficiaries, regardless of retention status (Figure 2, Table S2). By 2015, 43.5% of male beneficiaries were adherent, compared to 38.1% of female beneficiaries. Relative to non-Hispanic white beneficiaries (Figure 3), non-Hispanic Black beneficiaries had a higher prevalence of being not retained/not ART adherent (2002: 27.6% vs. 19.2%%; 2015: 18.3% vs. 15.1%) and a lower prevalence of being retained/ART adherent (2002: 19.4% vs. 25.7%; 2015: 27.4% vs. 35.7%). We further saw that non-Hispanic Black beneficiaries had a higher incidence of death across follow-up (2015: 29.8% vs. 25.0%), although the annual increase in the cumulative incidence of death became similar in the two groups by the end of the study period. Figure S1 and S2 visualize the sex- and race/ethnicity-stratified results when not considering incidence of death.

**Figure 2.**
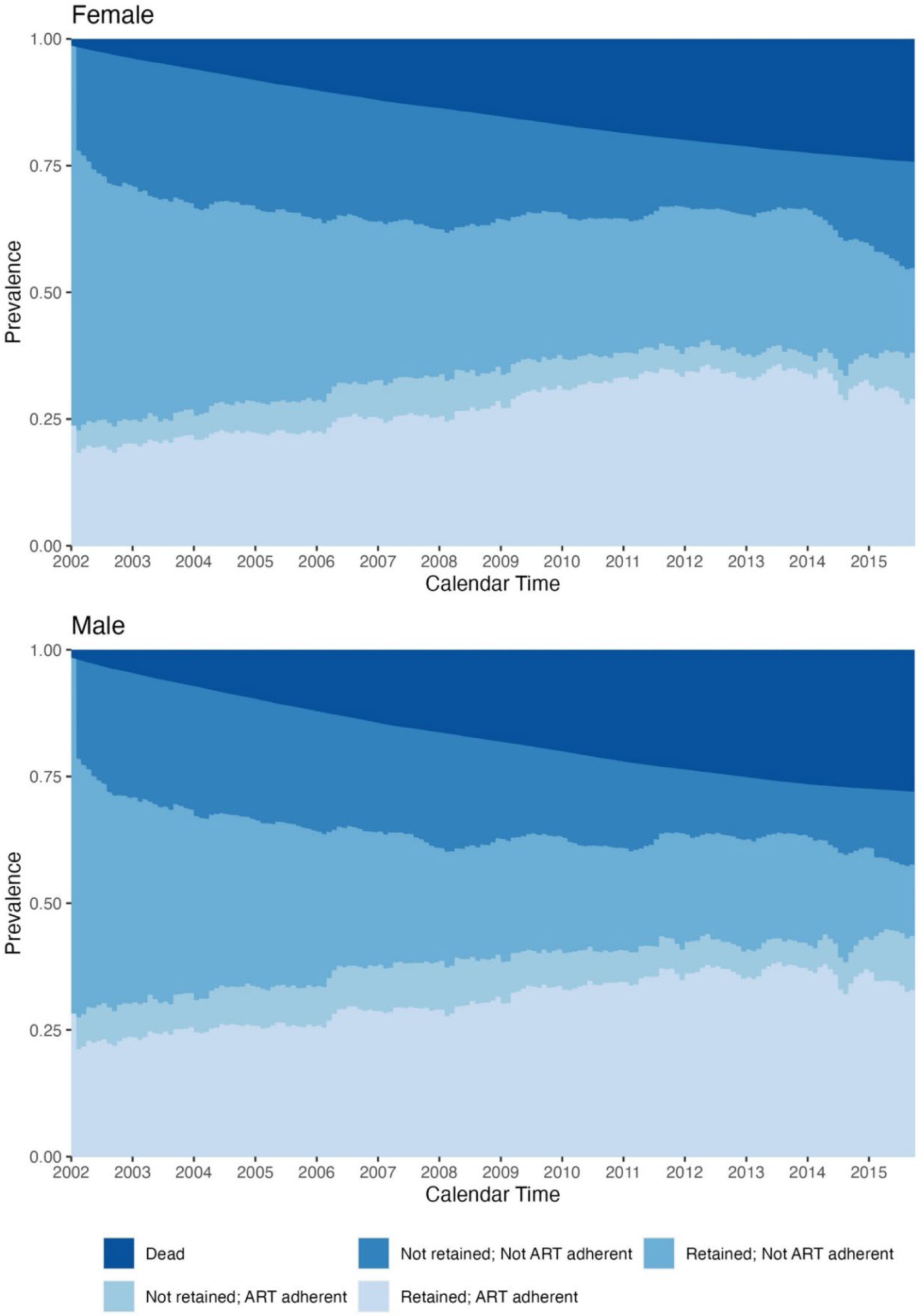
Weighted prevalence of HIV care cascade levels and incidence of death among male and female Medicaid beneficiaries, using an adherence level of 80%.

**Figure 3.**
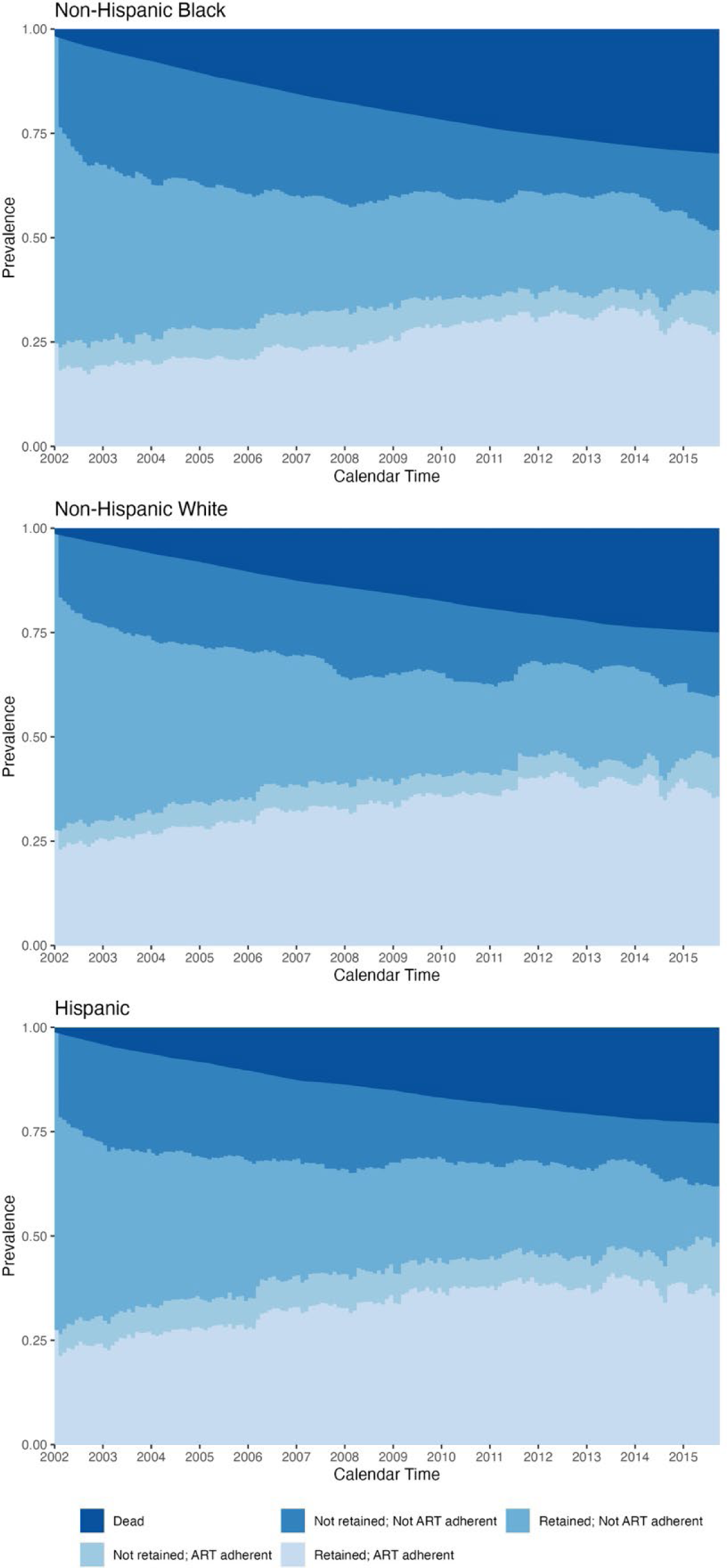
Weighted prevalence of HIV care cascade levels and incidence of death among non-Hispanic Black, non-Hispanic white, and Hispanic Medicaid beneficiaries, using an adherence level of 80.

The proportion retained/ART adherent was higher in all calendar years in the US West (2002: 26.6%; 2008: 45.2%; 2015: 48.2%) and Northeast (2002: 23.4%; 2008: 32.9%; 2015: 47.3%) regions than in the South (2002: 19.4%; 25.7%; 2015: 21.7%) or Midwest (2002: 17.3%; 2008: 19.1%; 2015: 16.2%) regions (Figure S3). We saw that the proportion retained/ART adherent was initially lower in the Midwest than in the South but then became higher across 2011-2013, although the trend reversed in 2014 and 2015. For the first half of the study period (2002-2009), the West had a lower prevalence of individuals not retained/not ART adherent than the Northeast (2002: 19.0% vs. 27.5%; 2009: 15.6% vs. 18.1%) and a higher prevalence of individuals retained/not ART adherent (2002: 47.3% vs. 39.7%; 2009: 26.9% vs. 22.3%). From 2010 onwards, though, the Northeast had a lower prevalence of individuals not retained/not ART adherent than the West and a similar or higher prevalence of individuals retained/not ART adherent. The Midwest initially had the highest prevalence of individuals who were retained/not ART adherent (2002: 56.3%), but the prevalence decreased over time; this region had the highest prevalence of individuals remaining in the not retained/not ART adherent state by 2015 (29.2%). Figure S4 visualizes the findings by region without considering death.

When ART adherence was defined based on any evidence of ART or an MPR ≥70% (Table S2 and S3, respectively), ART adherence was higher than in the main analysis. Under the MPR ≥70% definition, the proportion retained/ART adherent was 23.9% in 2002 and 33.2% in 2015. The opposite was true when we used a stricter definition for ART adherence. When the MPR cut-point was ≥90% (Table S4), that the prevalence of being retained/ART adherent (2002: 18.7%; 2013: 27.7%) and not retained/ART adherent (2002: 4.7%; 2013: 8.8%) was lower. We noted that the drop in prevalence of ART adherence was larger when the cut-point was shifted from 80% to 90% than when it was shifted from 70% to 80%. Nevertheless, trends were similar regardless of the MPR cutoff.

In the Appendix, we provide the crude results for the overall analyses (Tables S5-S8) and the results of the sensitivity analyses. When excluding CA from the analysis (Table S9), mortality incidence by 2015 was 3.7% higher than the main analysis, and the proportion retained/ART adherent was generally 1-3% lower than the main analysis. The difference in prevalence for all other states was generally <1% in all calendar years. When allowing all states to remain in the analysis until September 30, 2015 (Table S10), the proportion retained/ART adherent fell off starting in 2013 (1.4% lower); by the end of the 2014 and 2015, the prevalence was 5.2% and 6.8% lower than the main analysis. The prevalence of not retained/ART adherent was similarly lower.

## DISCUSSION

To our knowledge, our study represents the broadest examination of the HIV care cascade in Medicaid to date (Byrd et al., 2015; Health Resources and Services Administration, 2016). Across a time period that spanned the early highly active ART era to the early “Treat All” era, we saw an increase in the proportion of beneficiaries with HIV who were retained in care and ART adherent, overall and among all subgroups. By 2015, we saw that approximately half of beneficiaries still enrolled were retained in care (with 15% not ART adherent) and that 42% of beneficiaries were ART adherent (with 10% not retained in care). However, we saw meaningful disparities by race/ethnicity, with non-Hispanic Black beneficiaries being less likely to be retained and ART adherent than non-Hispanic white beneficiaries, and by US Census region, with the US South and Midwest trailing the West and Northeast. We present our findings in light of the competing event of death because this provides additional context for our results.

Mortality incidence among Medicaid beneficiaries with HIV was high during this time period (much higher than beneficiaries without HIV at similar ages (Rudolph et al., 2023)); furthermore, death represents an important negative consequence of less-than-ideal HIV care. Our findings for retention in care are similar to a 2010 CDC report, which found that 51% of PWH who were linked to care were retained in care (Centers for Disease Control and Prevention (CDC), 2011). This report found that 89% of PWH linked to care had been prescribed ART and 77% had achieved viral suppression. Even when not considering incidence of death (which would be more comparable to the CDC results), we estimated that 57.2% of Medicaid beneficiaries were adherent to ART in 2015. Our estimates of ART adherence are also lower than the proportion on ART and virally suppressed that have been reported in US-based HIV observational cohorts (Desir et al., 2019; Dombrowski et al., 2013; Lesko et al., 2017; Rebeiro et al., 2013). For example, the Center for AIDS Research Network of Integrated Clinical Systems reported that 89% of participants engaged in the study in 2010 were on ART and 70% had an undetectable viral load (Dombrowski et al., 2013). Similarly, the North American AIDS Cohort Collaboration on Research and Design reported that 75% of participants were consistently engaged in care between 2000-2008 (Rebeiro et al., 2013). In the same cohort, across participant’s first 5 years of enrollment, the estimates for proportion of time spent virally suppressed between 2000-2014 were similar to the prevalence of ART adherence estimated here (40.1% among Black participants and 44.8% among White participants) (Desir et al., 2019).

Multiple sources have reported that Black PWH in the US have worse retention in care and HIV treatment outcomes than other race groups (Althoff et al., 2014; Centers for Disease Control and Prevention, 2021; Desir et al., 2019; Hanna et al., 2013; Lillie-Blanton et al., 2010; Muthulingam et al., 2013; Rebeiro et al., 2013, 2017). We saw a similar disparity in Medicaid – a sample in which there should be comparable socioeconomic status due to the low-income eligibility requirement. That the disparity by race persists in Medicaid may reflect the additional structural and racism-related barriers that affect Black people in the US, as well as the geographic distribution of Black PWH (Michener, 2021). In our data, 40% of non-Hispanic Black beneficiaries lived in the US South, a larger proportion than any other racial/ethnic group, the region in which we observed the lowest prevalence of being retained in care/ART adherent and the highest mortality incidence. In 2016, the CDC similarly reported that PWH in the US South had a higher mortality rate than PWH in other regions (Centers for Disease Control and Prevention, 2016). Other studies have also reported lower retention in care in the South, relative to other regions (Rebeiro et al., 2016). In the context of Medicaid, states in the South tend to have higher barriers to remaining enrolled, and few Southern states have elected for Medicaid expansion, which has been associated with improvements in access to care and reductions in mortality and racial disparities (Guth et al., 2020; Guth & Published, 2021; “Medicaid Income Eligibility Limits for Adults as a Percent of the Federal Poverty Level,” n.d.).

Our analysis has several limitations worth noting. Medicaid claims data do not have direct information on HIV viral load or CD4 cell counts. We used ART adherence as a proxy for disease control, but it is likely that the proportion of beneficiaries who were suppressed differs from the proportion ART adherent as detected by claims. Our measure of ART adherence was based solely on prescription fills, not based on whether the supplied pills were taken. Given the way the medication possession ratio is calculated, though, one may reasonably assume that the pills were taken if an individual obtains a new prescription fill when the prior days supply ended. Our measure of ART adherence (and other HIV services) would also have been impacted if beneficiaries received ART from any source other than Medicaid, such as Ryan White. However, Ryan White funds only pay for prescriptions and services not covered by an individual’s insurance, and any beneficiary with full Medicaid benefits should have their ART prescriptions covered by Medicaid (HIV/AIDS Bureau, 2013).

We also examined only one definition for retention in HIV care. The definition requiring visits every 6 months aligns with the standard of care for people new to care or with uncontrolled HIV viremia (HHS Panel on Antiretroviral Guidelines for Adults & and Adolescents, 2024). It is possible that some PWH who were stably engaged in care may have needed to see their physician less frequently, e.g. once a year, and thus would have been flagged as “not retained in care.” In addition to being difficult to determine without HIV viral load data, a more relaxed retention definition would have majorly limited the analysis, given the median length of follow-up was 18 months. A further limitation was that, when examining the competing event of death, we lacked information on cause of death, so we could not distinguish whether beneficiaries were dying from AIDS or other causes.

Moreover, there are known issues with data quality in Medicaid that may differ by state. Unfortunately, our data mostly predate the DQ Atlas, which reports data quality by calendar year and state for Transformed Medicaid Statistical Information System Analytical Files (MACBIS, n.d.; Nguyen & Sanghavi, 2022). Potential data quality issues were why we excluded select states in select years. For the three states given earlier administrative censoring dates (NY, NC, MD), we noted in preliminary analyses a large drop-off in crude ART adherence that we could not pinpoint to any policy change. For example, the drop-off persisted when we excluded new enrollees post-Medicaid expansion in NY and MD. Almost no deaths were reported in the state of CA across the entire study period. We kept CA in the main analysis because the state was the second largest contributor of beneficiaries with HIV and because trends did not differ meaningfully in the sensitivity analysis excluding CA.

Finally, we made the decision to not allow beneficiaries to return to the analysis after a gap in Medicaid enrollment. While this limited the amount of follow-up, we would have no data on what happened to a beneficiary while they were not enrolled in Medicaid. Additionally, choosing to censor individuals when they disenrolled requires a weaker exchangeability assumption than allowing them to return (Robins et al., 1995).

To date, there has been a lack of knowledge on the state of the HIV care cascade among Medicaid beneficiaries, despite the fact that Medicaid represents a large and diverse sample of PWH (Roberts et al., 2023). Medicaid beneficiaries are a sample of low income and thus an underserved and vulnerable population of PWH, as our findings reinforce. Furthermore, this population has not been well captured by the existing clinical cohorts, which are often located at major centers of HIV care. In our sample, we saw that, despite being linked to care, less than half of beneficiaries with HIV were classified as ART adherent at any point in follow-up, indicating that many Medicaid beneficiaries with HIV were not virally suppressed during this time period. Future work ought to explore whether engagement with HIV care improved among Medicaid beneficiaries post-2015 and ought to investigate specific barriers to engagement in care among PWH insured by Medicaid. Overall, our findings highlight the importance of continuing to evaluate the HIV care cascade among Medicaid beneficiaries as a measure for HIV care.

## Data Availability

Data can be made available under a Data Use Agreement with the Centers for Medicare and Medicaid Services.

