## Supplemental Appendix for "Retention in care and antiretroviral therapy adherence among Medicaid beneficiaries with HIV, 2001-2015"

**Appendix Table of Contents.**


| Item | Description | Page |
|---|---|---|
| Table S1 | Codes used to define key variables | 3 |
| Table S2 | Weighted prevalence of each level of the HIV care cascade and cumulative incidence of mortality by end of the calendar year and the annual increase in the cumulative incidence of mortality by sex, using an adherence level of 80% | 4 |
| Figure S1 | Weighted prevalence of each level of the HIV care cascade by end of the calendar year not considering incidence of death by sex, using an adherence level of 80% | 5 |
| Table S3 | Weighted prevalence of each level of the HIV care cascade and cumulative incidence of mortality by end of the calendar year and the annual increase in the cumulative incidence of mortality by race/ethnicity, using an adherence level of 80% | 6 |
| Figure S2 | Weighted prevalence of each level of the HIV care cascade by end of the calendar year not considering incidence of death by race/ethnicity, using an adherence level of 80% | 8 |
| Figure S3 | Weighted prevalence of each level of the HIV care cascade and cumulative incidence of mortality by end of the calendar year by US Census Region, using an adherence level of 80% | 9 |
| Figure S4 | Weighted prevalence of each level of the HIV care cascade by end of the calendar year not considering incidence of death by US Census Region, using an adherence level of 80% | 10 |
| Table S4 | Weighted prevalence of each level of the HIV care cascade and cumulative incidence of mortality by end of the calendar year, using an adherence level of 0% | 11 |
| Table S5 | Weighted prevalence of each level of the HIV care cascade and cumulative incidence of mortality by end of the calendar year, using an adherence level of 70% | 12 |
| Table S6 | Weighted prevalence of each level of the HIV care cascade and cumulative incidence of mortality by end of the calendar year, using an adherence level of 90% | 13 |
| Table S7 | Crude prevalence of each level of the HIV care cascade and cumulative incidence of mortality by end of the calendar year, using an adherence level of 80% | 14 |

| | | |
|---|---|---|
| Table S8 | Crude prevalence of each level of the HIV care cascade and cumulative incidence of mortality by end of the calendar year, using an adherence level of 0% | 15 |
| Table S9 | Crude prevalence of each level of the HIV care cascade and cumulative incidence of mortality by end of the calendar year, using an adherence level of 70% | 16 |
| Table S10 | Crude prevalence of each level of the HIV care cascade and cumulative incidence of mortality by end of the calendar year, using an adherence level of 90% | 17 |
| Table S11 | Weighted prevalence of each level of the HIV care cascade and cumulative incidence of mortality by end of the calendar year, using an adherence level of 80% and excluding California | 18 |
| Table S12 | Weighted prevalence of each level of the HIV care cascade and cumulative incidence of mortality by end of the calendar year, using an adherence level of 80% and using September 30, 2015 as the administrative censoring date for all states | 19 |

Table S1. Codes used to define key variables[a]

| Variable | CPT/HCPCS | ICD-9 |
| --- | --- | --- |
| HIV Diagnosis | --- | 042, 079.53, 795.71, V08 |
| Office visits[b] | 99201, 99202, 99203, 99204, 99205, 99212, 99213, 99214, 99215 | --- |
| CD4/CD8 measurements | 86359, 86360, 86361 | --- |
| Viral load tests | 87534, 87535, 87536, 87537, 87538, 87539 | --- |
| Myocardial infarction | --- | 410.X, 412.X |
| Congestive heart failure | --- | 398.91, 402.01, 402.11, 402.91, 404.01, 404.03, 404.11, 404.13, 404.91, 404.93, 425.4-425.9, 428.X |
| Peripheral vascular disease | --- | 093.0, 437.3, 440.X, 441.X, 443.1-443.9, 447.1, 557.1, 557.9, V43.4 |
| Cerebrovascular disease | --- | 362.34, 430.X-438.X |
| Dementia | --- | 290.X, 294.1, 331.2 |
| Chronic pulmonary disease | --- | 416.8, 416.9, 490.X-505.X, 506.4, 508.1, 508.8 |
| Rheumatic disease | --- | 446.5, 710.0-710.4, 714.0-714.2, 714.8, 725.X |
| Peptic ulcer disease | --- | 531.X-534.X |
| Mild liver disease | --- | 070.22, 070.23, 070.32, 070.33, 070.44, 070.54, 070.6, 070.9, 570.X, 571.X, 573.3, 573.4, 573.8, 573.9, V42.7 |
| Moderate/severe liver disease | | 456.0-456.2, 572.2-572.8 |
| Diabetes without chronic complication | --- | 250.0-250.3, 250.8, 250.9 |
| Diabetes with chronic complication | --- | 250.4-250.7 |
| Hemiplegia/paraplegia | --- | 334.1, 342.X, 343.X, 344.0-344.6, 344.9 |
| Renal disease | --- | 403.01, 403.11, 403.91, 404.02, 404.03, 404.12, 404.13, 404.92, 404.93, 582.X, 583.0-583.7, 585.X, 586.X, 588.0, V42.0, V45.1, V56.X |
| Malignancy | --- | 140.X-172.X, 174.X-195.8, 200.X-208.X, 238.6 |
| Metastatic solid tumor | --- | 196.X-199.X |

Abbreviations: CPT, current procedural terminology; HCPCS, healthcare common procedure coding system; ICD, international classification of diseases

[a]See supplemental file ndc_class.txt for NDC codes used to identify antiretroviral therapy

[b]Where revenue center code not in 0450-0459, 0981, 0516, 0526

Table S2. Weighted prevalence of each level of the HIV care cascade and cumulative incidence of mortality by end of the calendar year and the annual increase in the cumulative incidence of mortality by sex, using an adherence level of 80%

| Calendar Year | Not retained Not ART adherent | | Retained Not ART adherent | | Not retained ART adherent | | Retained ART adherent | | Mortality Incidence | | Annual Increase in Mortality | |
|---|---|---|---|---|---|---|---|---|---|---|---|---|
| | % | 95% CI | % | 95% CI | % | 95% CI | % | 95% CI | % | 95% CI | % | 95% CI |
| *Female beneficiaries* | | | | | | | | | | | | |
| 2002 | 25.0 | (24.5, 25.6) | 46.4 | (45.9, 47.0) | 4.5 | (4.2, 4.7) | 20.2 | (19.8, 20.6) | 3.9 | (3.7, 4.1) | 2.5 | (2.3, 2.8) |
| 2003 | 26.5 | (26.0, 27.1) | 40.7 | (40.2, 41.3) | 5.0 | (4.8, 5.3) | 21.8 | (21.4, 22.3) | 5.9 | (5.7, 6.2) | 2.1 | (1.7, 2.4) |
| 2004 | 24.8 | (24.3, 25.3) | 38.5 | (38.0, 39.1) | 6.1 | (5.8, 6.4) | 22.4 | (21.9, 22.8) | 8.1 | (7.8, 8.5) | 2.2 | (1.8, 2.6) |
| 2005 | 25.4 | (24.9, 25.9) | 35.9 | (35.3, 36.4) | 6.3 | (6.0, 6.6) | 22.2 | (21.8, 22.7) | 10.1 | (9.8, 10.5) | 2.0 | (1.5, 2.5) |
| 2006 | 24.0 | (23.5, 24.5) | 31.3 | (30.7, 31.8) | 7.2 | (6.9, 7.5) | 25.4 | (24.9, 25.9) | 12.1 | (11.7, 12.5) | 1.9 | (1.4, 2.4) |
| 2007 | 23.9 | (23.4, 24.5) | 29.1 | (28.5, 29.6) | 8.0 | (7.8, 8.4) | 25.3 | (24.8, 25.8) | 13.6 | (13.3, 14.0) | 1.6 | (1.0, 2.1) |
| 2008 | 20.2 | (19.7, 20.7) | 29.2 | (28.6, 29.7) | 6.8 | (6.5, 7.1) | 28.5 | (28.0, 29.1) | 15.3 | (14.9, 15.7) | 1.7 | (1.1, 2.2) |
| 2009 | 17.3 | (16.8, 17.7) | 28.1 | (27.5, 28.6) | 6.0 | (5.7, 6.3) | 31.6 | (31.0, 32.2) | 17.0 | (16.6, 17.5) | 1.7 | (1.1, 2.3) |
| 2010 | 16.9 | (16.4, 17.3) | 26.5 | (26.0, 27.0) | 4.8 | (4.6, 5.1) | 33.2 | (32.6, 33.8) | 18.6 | (18.1, 19.0) | 1.6 | (0.9, 2.2) |
| 2011 | 13.2 | (12.8, 13.6) | 29.1 | (28.5, 29.6) | 4.5 | (4.2, 4.8) | 33.3 | (32.7, 33.9) | 19.9 | (19.5, 20.4) | 1.4 | (0.7, 2.0) |
| 2012 | 13.5 | (13.0, 13.9) | 27.8 | (27.3, 28.4) | 4.5 | (4.3, 4.8) | 33.0 | (32.4, 33.5) | 21.2 | (20.7, 21.7) | 1.3 | (0.6, 1.9) |
| 2013 | 11.0 | (10.6, 11.4) | 28.9 | (28.4, 29.4) | 3.7 | (3.5, 3.9) | 34.0 | (33.4, 34.5) | 22.4 | (21.9, 22.9) | 1.2 | (0.5, 1.9) |
| 2014 | 16.7 | (16.1, 17.3) | 21.9 | (21.2, 22.5) | 5.2 | (4.9, 5.6) | 32.7 | (32.0, 33.3) | 23.5 | (23.0, 24.0) | 1.1 | (0.4, 1.7) |
| 2015 | 21.0 | (20.3, 21.7) | 16.7 | (16.2, 17.2) | 9.1 | (8.7, 9.6) | 29.0 | (28.4, 29.6) | 24.2 | (23.8, 24.8) | 0.8 | (0.1, 1.5) |
| *Male beneficiaries* | | | | | | | | | | | | |
| 2002 | 24.7 | (24.2, 25.1) | 40.6 | (40.1, 41.1) | 6.5 | (6.3, 6.8) | 23.6 | (23.2, 24.0) | 4.6 | (4.3, 4.8) | 3.0 | (2.7, 3.3) |
| 2003 | 24.4 | (23.9, 24.9) | 36.2 | (35.7, 36.7) | 6.7 | (6.4, 6.9) | 25.6 | (25.1, 26.0) | 7.2 | (6.9, 7.5) | 2.6 | (2.2, 3.0) |
| 2004 | 23.8 | (23.3, 24.3) | 32.9 | (32.4, 33.3) | 7.9 | (7.6, 8.2) | 25.8 | (25.4, 26.3) | 9.7 | (9.3, 10.0) | 2.5 | (2.1, 2.9) |
| 2005 | 23.6 | (23.2, 24.1) | 30.9 | (30.3, 31.3) | 7.7 | (7.5, 8.0) | 25.7 | (25.2, 26.2) | 12.1 | (11.7, 12.5) | 2.5 | (2.0, 2.9) |
| 2006 | 21.7 | (21.2, 22.1) | 26.4 | (25.9, 26.8) | 8.8 | (8.6, 9.2) | 28.7 | (28.2, 29.2) | 14.4 | (14.0, 14.8) | 2.3 | (1.8, 2.8) |
| 2007 | 22.8 | (22.4, 23.3) | 22.6 | (22.2, 23.0) | 9.4 | (9.1, 9.7) | 29.0 | (28.4, 29.4) | 16.3 | (15.9, 16.7) | 1.9 | (1.3, 2.4) |
| 2008 | 19.1 | (18.6, 19.6) | 22.9 | (22.5, 23.4) | 8.3 | (8.0, 8.6) | 31.5 | (31.1, 32.0) | 18.1 | (17.7, 18.6) | 1.9 | (1.3, 2.5) |
| 2009 | 16.8 | (16.3, 17.2) | 22.3 | (21.9, 22.8) | 7.2 | (7.0, 7.5) | 33.7 | (33.2, 34.2) | 20.0 | (19.6, 20.5) | 1.9 | (1.2, 2.5) |
| 2010 | 17.1 | (16.7, 17.5) | 20.1 | (19.7, 20.5) | 6.3 | (6.1, 6.6) | 34.4 | (33.9, 34.9) | 22.0 | (21.6, 22.5) | 2.0 | (1.4, 2.7) |
| 2011 | 12.6 | (12.3, 13.0) | 22.9 | (22.5, 23.4) | 6.0 | (5.8, 6.3) | 34.8 | (34.3, 35.3) | 23.6 | (23.1, 24.1) | 1.6 | (0.9, 2.2) |
| 2012 | 12.4 | (12.1, 12.8) | 22.0 | (21.6, 22.4) | 5.4 | (5.2, 5.6) | 35.0 | (34.6, 35.6) | 25.1 | (24.6, 25.6) | 1.5 | (0.8, 2.2) |
| 2013 | 10.1 | (9.7, 10.4) | 21.2 | (20.8, 21.6) | 5.0 | (4.8, 5.2) | 37.2 | (36.7, 37.7) | 26.5 | (26.1, 27.0) | 1.4 | (0.8, 2.1) |
| 2014 | 11.6 | (11.2, 12.0) | 17.7 | (17.3, 18.1) | 6.3 | (6.0, 6.6) | 37.0 | (36.4, 37.5) | 27.4 | (26.9, 27.9) | 0.9 | (0.2, 1.6) |
| 2015 | 14.4 | (14.0, 14.8) | 14.1 | (13.8, 14.4) | 10.6 | (10.3, 11.0) | 32.9 | (32.4, 33.3) | 28.0 | (27.5, 28.5) | 0.6 | (0.0, 1.3) |

Abbreviations: ART, antiretroviral therapy; CI, confidence interval

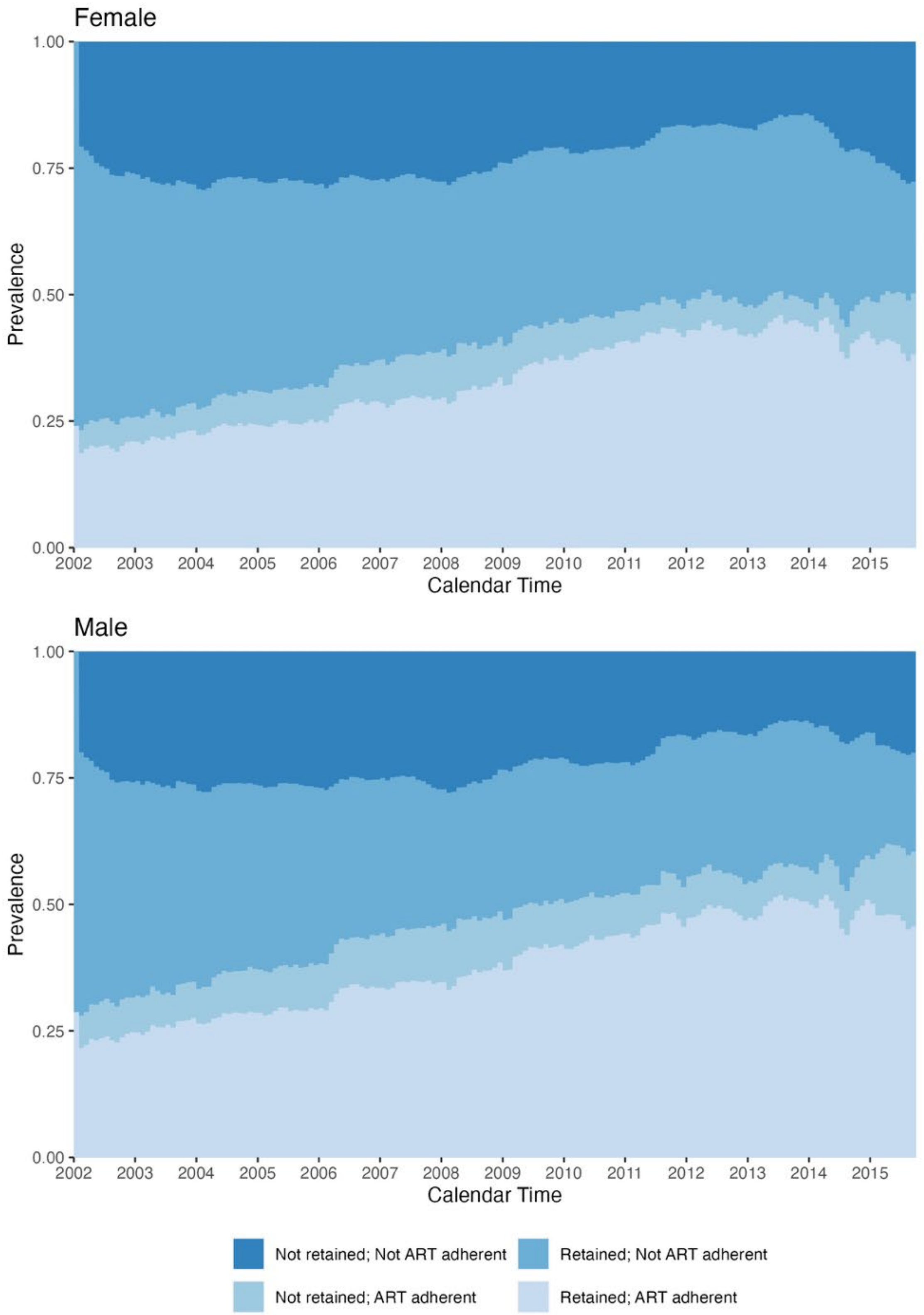


Figure S1. Weighted prevalence of each level of the HIV care cascade by end of the calendar year not considering incidence of death by sex, using an adherence level of 80%

Table S3. Weighted prevalence of each level of the HIV care cascade and cumulative incidence of mortality by end of the calendar year and the annual increase in the cumulative incidence of mortality by race/ethnicity, using an adherence level of 80%

| Calendar Year | Not retained Not ART adherent | | Retained Not ART adherent | | Not retained ART adherent | | Retained ART adherent | | Mortality Incidence | | Annual Increase in Mortality | |
|---|---|---|---|---|---|---|---|---|---|---|---|---|
| | % | 95% CI | % | 95% CI | % | 95% CI | % | 95% CI | % | 95% CI | % | 95% CI |
| *Non-Hispanic Black beneficiaries* | | | | | | | | | | | | |
| 2002 | 27.6 | (27.1, 28.2) | 42.4 | (41.8, 42.9) | 5.6 | (5.3, 5.9) | 19.4 | (19.0, 19.8) | 5.0 | (4.8, 5.3) | 3.3 | (3.0, 3.6) |
| 2003 | 28.4 | (27.9, 28.9) | 37.4 | (36.9, 37.9) | 6.1 | (5.8, 6.3) | 20.5 | (20.2, 21.0) | 7.6 | (7.4, 7.9) | 2.6 | (2.3, 3.0) |
| 2004 | 26.3 | (25.8, 26.8) | 34.7 | (34.2, 35.2) | 7.3 | (7.0, 7.6) | 21.1 | (20.7, 21.5) | 10.5 | (10.2, 10.9) | 2.8 | (2.4, 3.3) |
| 2005 | 26.6 | (26.1, 27.1) | 32.4 | (31.9, 32.8) | 7.3 | (7.0, 7.6) | 20.7 | (20.3, 21.1) | 13.0 | (12.7, 13.4) | 2.6 | (2.1, 3.1) |
| 2006 | 24.6 | (24.2, 25.1) | 28.1 | (27.6, 28.6) | 8.2 | (7.9, 8.5) | 23.6 | (23.1, 24.0) | 15.5 | (15.1, 15.9) | 2.5 | (1.9, 3.0) |
| 2007 | 24.4 | (23.9, 24.8) | 25.3 | (24.9, 25.7) | 9.0 | (8.7, 9.3) | 23.7 | (23.2, 24.1) | 17.6 | (17.2, 18.1) | 2.1 | (1.5, 2.7) |
| 2008 | 20.2 | (19.7, 20.7) | 25.7 | (25.3, 26.2) | 7.9 | (7.6, 8.2) | 26.5 | (26.0, 26.9) | 19.7 | (19.3, 20.2) | 2.1 | (1.5, 2.7) |
| 2009 | 17.3 | (16.9, 17.8) | 25.0 | (24.6, 25.4) | 6.7 | (6.4, 6.9) | 29.3 | (28.8, 29.8) | 21.7 | (21.3, 22.2) | 2.0 | (1.4, 2.6) |
| 2010 | 17.5 | (17.0, 17.9) | 22.7 | (22.3, 23.2) | 5.7 | (5.4, 6.0) | 30.4 | (30.0, 30.9) | 23.7 | (23.3, 24.2) | 2.0 | (1.3, 2.6) |
| 2011 | 13.9 | (13.5, 14.3) | 25.5 | (25.1, 26.0) | 5.5 | (5.3, 5.8) | 29.8 | (29.3, 30.2) | 25.3 | (24.8, 25.7) | 1.5 | (0.9, 2.2) |
| 2012 | 13.7 | (13.4, 14.1) | 24.0 | (23.5, 24.4) | 5.2 | (5.0, 5.4) | 30.4 | (29.9, 30.9) | 26.7 | (26.3, 27.2) | 1.4 | (0.8, 2.1) |
| 2013 | 11.3 | (10.9, 11.6) | 23.3 | (22.9, 23.7) | 4.5 | (4.3, 4.7) | 32.9 | (32.5, 33.4) | 28.0 | (27.6, 28.6) | 1.4 | (0.7, 2.0) |
| 2014 | 14.4 | (13.9, 14.8) | 19.6 | (19.1, 20.0) | 6.0 | (5.7, 6.3) | 30.9 | (30.4, 31.5) | 29.2 | (28.7, 29.6) | 1.1 | (0.4, 1.8) |
| 2015 | 18.3 | (17.9, 18.8) | 14.6 | (14.1, 15.0) | 9.9 | (9.5, 10.3) | 27.4 | (26.8, 27.9) | 29.8 | (29.4, 30.3) | 0.7 | (0.0, 1.3) |
| *Non-Hispanic white beneficiaries* | | | | | | | | | | | | |
| 2002 | 19.2 | (18.5, 19.9) | 47.0 | (46.1, 47.8) | 4.3 | (4.0, 4.7) | 25.7 | (24.9, 26.4) | 3.8 | (3.4, 4.1) | 2.4 | (2.0, 2.8) |
| 2003 | 19.8 | (19.1, 20.5) | 41.7 | (40.9, 42.5) | 5.0 | (4.6, 5.4) | 27.4 | (26.7, 28.2) | 6.1 | (5.6, 6.4) | 2.3 | (1.7, 2.8) |
| 2004 | 20.0 | (19.4, 20.7) | 37.6 | (36.7, 38.4) | 5.9 | (5.5, 6.3) | 28.5 | (27.8, 29.2) | 8.1 | (7.6, 8.5) | 2.0 | (1.4, 2.6) |
| 2005 | 19.1 | (18.5, 19.8) | 35.5 | (34.7, 36.3) | 5.2 | (4.8, 5.6) | 29.7 | (28.9, 30.4) | 10.4 | (10.0, 11.0) | 2.4 | (1.7, 3.1) |
| 2006 | 17.9 | (17.2, 18.6) | 31.1 | (30.4, 31.9) | 6.3 | (5.9, 6.7) | 32.1 | (31.3, 32.9) | 12.6 | (12.0, 13.2) | 2.2 | (1.4, 2.9) |
| 2007 | 21.5 | (20.7, 22.3) | 25.7 | (24.9, 26.5) | 6.0 | (5.6, 6.4) | 32.7 | (31.8, 33.5) | 14.1 | (13.5, 14.7) | 1.5 | (0.7, 2.3) |
| 2008 | 19.0 | (18.3, 19.8) | 25.3 | (24.6, 26.1) | 5.5 | (5.1, 5.9) | 34.4 | (33.6, 35.2) | 15.7 | (15.2, 16.4) | 1.6 | (0.8, 2.5) |
| 2009 | 17.3 | (16.5, 18.2) | 24.2 | (23.4, 25.1) | 4.7 | (4.3, 5.1) | 36.2 | (35.4, 37.0) | 17.4 | (16.8, 18.0) | 1.7 | (0.8, 2.5) |
| 2010 | 18.0 | (17.2, 18.7) | 21.4 | (20.7, 22.2) | 5.1 | (4.7, 5.5) | 36.2 | (35.3, 37.0) | 19.3 | (18.7, 20.0) | 1.9 | (1.0, 2.8) |
| 2011 | 11.1 | (10.5, 11.8) | 23.1 | (22.3, 23.8) | 5.0 | (4.6, 5.4) | 40.1 | (39.2, 40.9) | 20.7 | (20.0, 21.5) | 1.4 | (0.4, 2.4) |
| 2012 | 11.6 | (10.9, 12.2) | 24.0 | (23.3, 24.8) | 4.7 | (4.3, 5.1) | 37.5 | (36.5, 38.3) | 22.3 | (21.5, 23.0) | 1.5 | (0.5, 2.6) |
| 2013 | 9.5 | (8.9, 10.1) | 24.5 | (23.7, 25.3) | 4.0 | (3.6, 4.3) | 38.4 | (37.6, 39.2) | 23.7 | (23.0, 24.5) | 1.4 | (0.4, 2.5) |
| 2014 | 12.7 | (12.1, 13.5) | 18.0 | (17.4, 18.7) | 4.9 | (4.4, 5.3) | 40.0 | (39.0, 40.7) | 24.4 | (23.7, 25.2) | 0.7 | (0.0, 1.8) |
| 2015 | 15.1 | (14.4, 15.8) | 14.8 | (14.3, 15.5) | 9.4 | (8.8, 9.9) | 35.7 | (34.8, 36.4) | 25.0 | (24.3, 25.8) | 0.6 | (0.0, 1.6) |
| *Hispanic beneficiaries* | | | | | | | | | | | | |
| 2002 | 23.3 | (22.3, 24.2) | 41.7 | (40.6, 42.8) | 6.7 | (6.1, 7.3) | 24.2 | (23.1, 25.2) | 4.2 | (3.7, 4.7) | 3.0 | (2.4, 3.6) |
| 2003 | 22.9 | (21.8, 23.9) | 37.1 | (35.8, 38.2) | 6.7 | (6.0, 7.4) | 27.0 | (25.9, 28.2) | 6.3 | (5.8, 7.0) | 2.1 | (1.4, 2.9) |
| 2004 | 22.6 | (21.5, 23.8) | 33.5 | (32.4, 34.8) | 7.4 | (6.7, 8.0) | 28.2 | (27.1, 29.3) | 8.3 | (7.7, 9.0) | 2.0 | (1.1, 2.9) |
| 2005 | 21.4 | (20.2, 22.5) | 33.8 | (32.6, 35.1) | 6.8 | (6.2, 7.5) | 27.6 | (26.4, 28.7) | 10.4 | (9.7, 11.1) | 2.1 | (1.1, 3.0) |
| 2006 | 19.1 | (17.9, 20.3) | 27.9 | (26.6, 29.1) | 7.4 | (6.7, 8.1) | 32.9 | (31.7, 34.1) | 12.7 | (11.9, 13.5) | 2.3 | (1.2, 3.4) |

| | | | | | | | | | | | | |
|---|---|---|---|---|---|---|---|---|---|---|---|---|
| 2007 | 20.2 | (19.0, 21.4) | 25.3 | (24.2, 26.5) | 8.2 | (7.4, 8.9) | 32.7 | (31.4, 33.9) | 13.7 | (12.8, 14.5) | 1.1 | (0.0, 2.2) |
| 2008 | 17.4 | (16.1, 18.4) | 24.4 | (23.3, 25.6) | 7.3 | (6.6, 8.1) | 35.8 | (34.6, 37.2) | 15.0 | (14.2, 15.9) | 1.3 | (0.1, 2.5) |
| 2009 | 14.2 | (13.1, 15.3) | 24.2 | (23.1, 25.5) | 7.2 | (6.4, 8.0) | 37.5 | (36.2, 38.8) | 16.9 | (15.9, 17.8) | 1.8 | (0.5, 3.1) |
| 2010 | 14.1 | (13.1, 15.2) | 23.3 | (22.2, 24.4) | 6.9 | (6.1, 7.6) | 37.6 | (36.2, 38.8) | 18.2 | (17.2, 19.2) | 1.3 | (0.0, 2.7) |
| 2011 | 12.5 | (11.6, 13.5) | 22.4 | (21.4, 23.6) | 6.8 | (6.1, 7.6) | 38.8 | (37.4, 40.0) | 19.5 | (18.5, 20.6) | 1.3 | (0.0, 2.8) |
| 2012 | 13.4 | (12.5, 14.4) | 21.5 | (20.3, 22.6) | 7.2 | (6.4, 7.8) | 37.2 | (35.8, 38.7) | 20.7 | (19.7, 21.8) | 1.3 | (0.0, 2.7) |
| 2013 | 10.1 | (9.2, 11.2) | 21.4 | (20.2, 22.5) | 6.9 | (6.2, 7.6) | 39.7 | (38.4, 41.0) | 21.9 | (20.9, 23.0) | 1.2 | (0.0, 2.7) |
| 2014 | 13.4 | (12.4, 14.5) | 17.0 | (16.1, 18.0) | 8.1 | (7.2, 8.9) | 38.9 | (37.6, 40.1) | 22.6 | (21.6, 23.7) | 0.6 | (0.0, 2.1) |
| 2015 | 15.1 | (14.0, 16.1) | 13.5 | (12.8, 14.3) | 12.0 | (11.2, 12.9) | 36.4 | (35.2, 37.5) | 23.0 | (22.0, 24.2) | 0.5 | (0.0, 2.0) |

Abbreviations: ART, antiretroviral therapy; CI, confidence interval

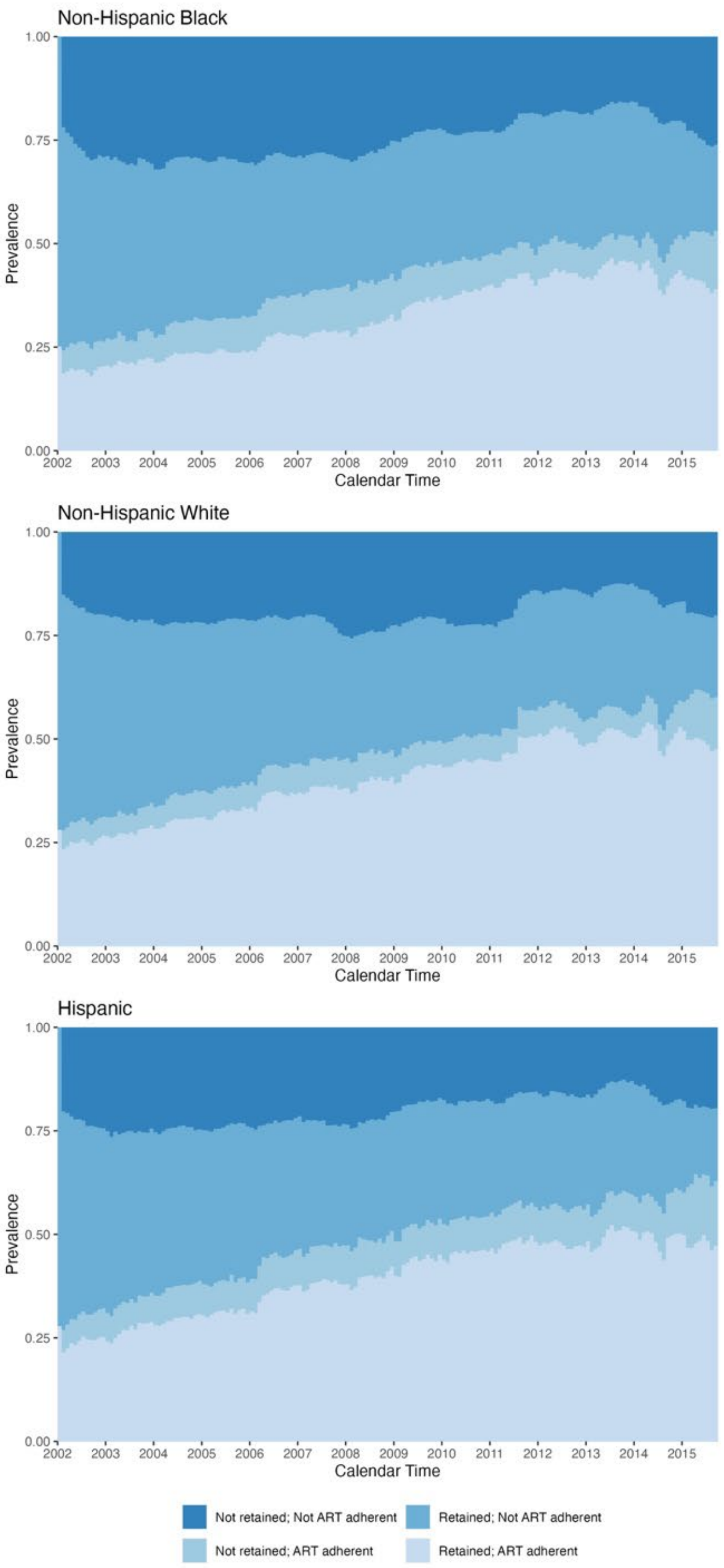


Figure S2. Weighted prevalence of each level of the HIV care cascade by end of the calendar year not considering incidence of death by race/ethnicity, using an adherence level of 80%

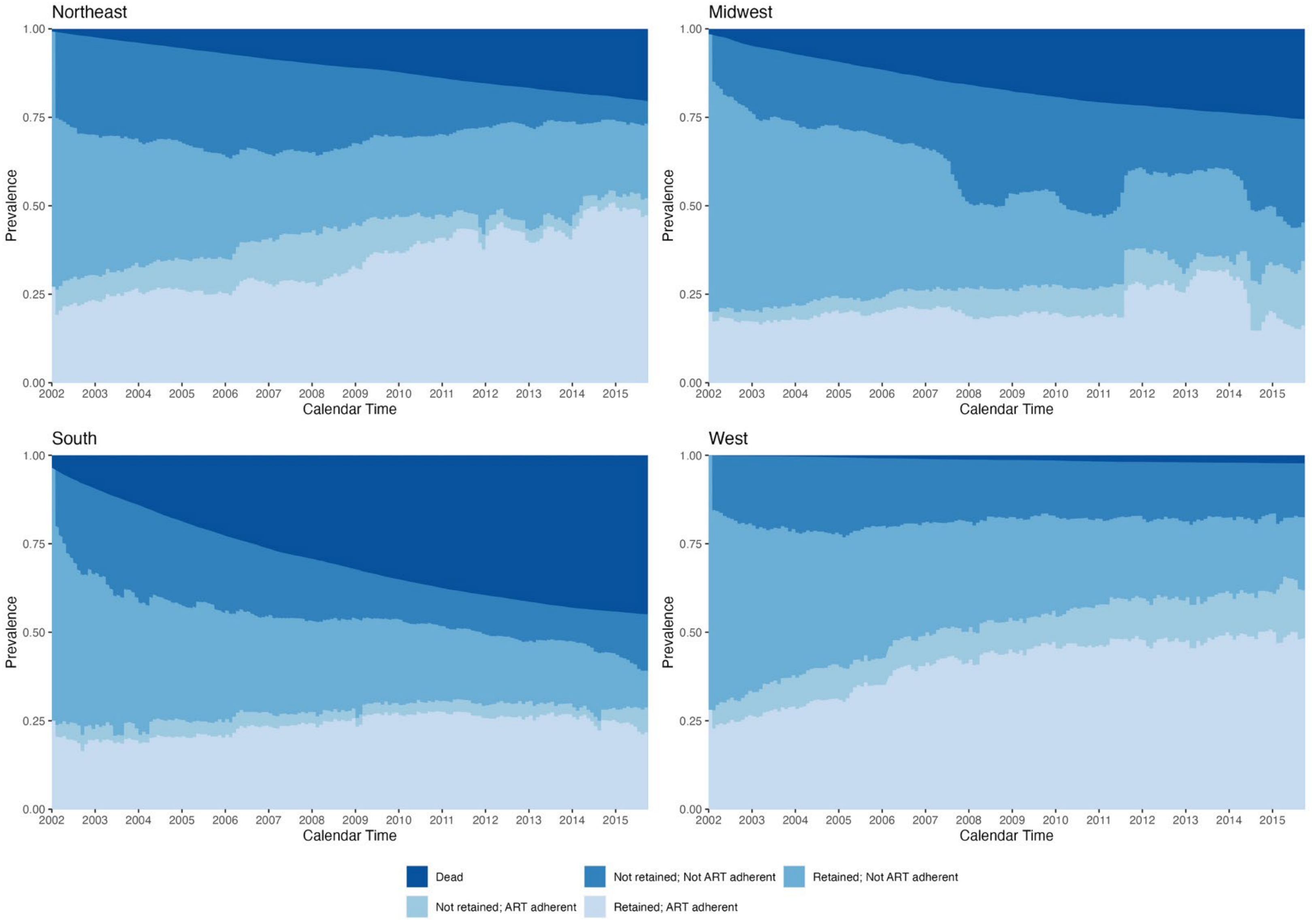


Figure S3. Weighted prevalence of each level of the HIV care cascade and cumulative incidence of mortality by end of the calendar year by US Census Region, using an adherence level of 80%

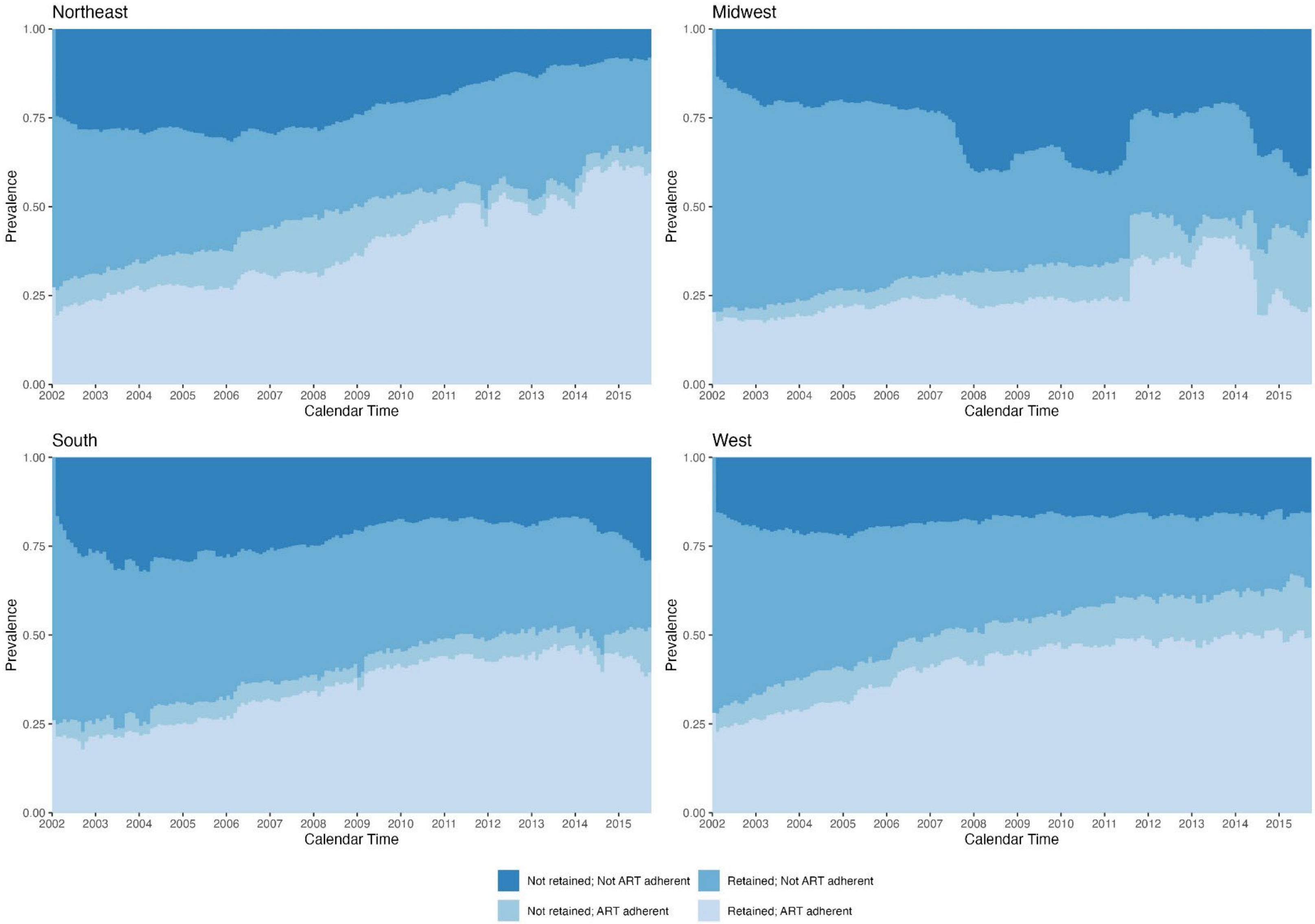


Figure S4. Weighted prevalence of each level of the HIV care cascade by end of the calendar year not considering incidence of death by US Census Region, using an adherence level of 80%

Table S4. Weighted prevalence of each level of the HIV care cascade and cumulative incidence of mortality by end of the calendar year, using an adherence level of 0%

| Calendar Year | Not retained Not ART adherent | | Retained Not ART adherent | | Not retained ART adherent | | Retained ART adherent | | Mortality Incidence | |
|---|---|---|---|---|---|---|---|---|---|---|
| | % | 95% CI | % | 95% CI | % | 95% CI | % | 95% CI | % | 95% CI |
| 2002 | 21.9 | (21.6, 22.3) | 34.4 | (34.1, 34.8) | 8.4 | (8.2, 8.6) | 31.0 | (30.6, 31.4) | 4.3 | (4.1, 4.4) |
| 2003 | 22.7 | (22.3, 23.0) | 30.1 | (29.7, 30.4) | 8.5 | (8.3, 8.8) | 32.2 | (31.8, 32.5) | 6.6 | (6.4, 6.8) |
| 2004 | 21.7 | (21.4, 22.0) | 27.5 | (27.2, 27.9) | 9.6 | (9.3, 9.8) | 32.2 | (31.8, 32.5) | 9.0 | (8.7, 9.2) |
| 2005 | 21.9 | (21.5, 22.2) | 25.5 | (25.2, 25.8) | 9.7 | (9.5, 9.9) | 31.7 | (31.4, 32.1) | 11.2 | (11.0, 11.5) |
| 2006 | 20.0 | (19.7, 20.4) | 20.5 | (20.3, 20.9) | 10.9 | (10.6, 11.1) | 35.2 | (34.8, 35.6) | 13.4 | (13.1, 13.6) |
| 2007 | 20.8 | (20.4, 21.1) | 17.7 | (17.4, 17.9) | 11.4 | (11.2, 11.7) | 35.0 | (34.7, 35.4) | 15.1 | (14.9, 15.4) |
| 2008 | 17.6 | (17.3, 18.0) | 17.9 | (17.5, 18.2) | 9.7 | (9.5, 9.9) | 37.9 | (37.5, 38.3) | 16.9 | (16.6, 17.2) |
| 2009 | 15.3 | (15.0, 15.6) | 16.7 | (16.4, 17.0) | 8.6 | (8.4, 8.8) | 40.7 | (40.3, 41.1) | 18.7 | (18.4, 19.0) |
| 2010 | 15.4 | (15.0, 15.7) | 14.3 | (14.0, 14.6) | 7.5 | (7.3, 7.7) | 42.3 | (41.9, 42.7) | 20.6 | (20.2, 20.9) |
| 2011 | 11.3 | (11.0, 11.5) | 15.8 | (15.5, 16.1) | 7.1 | (6.9, 7.3) | 43.8 | (43.4, 44.2) | 22.0 | (21.7, 22.4) |
| 2012 | 10.8 | (10.6, 11.0) | 13.8 | (13.6, 14.1) | 7.1 | (6.9, 7.4) | 44.8 | (44.4, 45.2) | 23.5 | (23.1, 23.8) |
| 2013 | 8.9 | (8.6, 9.1) | 14.9 | (14.6, 15.1) | 6.0 | (5.8, 6.1) | 45.5 | (45.1, 45.9) | 24.8 | (24.5, 25.2) |
| 2014 | 12.0 | (11.7, 12.3) | 11.5 | (11.2, 11.8) | 7.7 | (7.5, 8.0) | 43.0 | (42.5, 43.5) | 25.7 | (25.4, 26.1) |
| 2015 | 14.2 | (13.9, 14.5) | 8.2 | (8.0, 8.5) | 13.2 | (12.9, 13.5) | 38.0 | (37.5, 38.4) | 26.4 | (26.0, 26.8) |

Abbreviations: ART, antiretroviral therapy; CI, confidence interval

Table S5. Weighted prevalence of each level of the HIV care cascade and cumulative incidence of mortality by end of the calendar year, using an adherence level of 70%

| Calendar Year | Not retained Not ART adherent | | Retained Not ART adherent | | Not retained ART adherent | | Retained ART adherent | | Mortality Incidence | |
|---|---|---|---|---|---|---|---|---|---|---|
| | % | 95% CI | % | 95% CI | % | 95% CI | % | 95% CI | % | 95% CI |
| 2002 | 24.3 | (24.0, 24.6) | 41.4 | (41.1, 41.8) | 6.1 | (5.9, 6.3) | 23.9 | (23.6, 24.3) | 4.3 | (4.1, 4.4) |
| 2003 | 24.9 | (24.5, 25.2) | 36.4 | (36.0, 36.8) | 6.4 | (6.2, 6.6) | 25.7 | (25.4, 26.0) | 6.6 | (6.4, 6.8) |
| 2004 | 23.6 | (23.3, 24.0) | 33.3 | (33.0, 33.7) | 7.7 | (7.5, 7.9) | 26.4 | (26.0, 26.7) | 9.0 | (8.8, 9.2) |
| 2005 | 23.8 | (23.4, 24.1) | 31.0 | (30.7, 31.4) | 7.8 | (7.5, 8.0) | 26.2 | (25.9, 26.6) | 11.2 | (11.0, 11.5) |
| 2006 | 21.9 | (21.6, 22.3) | 26.2 | (25.9, 26.6) | 8.9 | (8.7, 9.1) | 29.6 | (29.2, 29.9) | 13.4 | (13.1, 13.7) |
| 2007 | 22.6 | (22.2, 22.9) | 23.2 | (22.8, 23.5) | 9.6 | (9.3, 9.8) | 29.6 | (29.2, 29.9) | 15.1 | (14.9, 15.4) |
| 2008 | 19.1 | (18.8, 19.5) | 23.7 | (23.4, 24.1) | 8.1 | (7.9, 8.3) | 32.1 | (31.8, 32.5) | 16.9 | (16.6, 17.2) |
| 2009 | 16.6 | (16.2, 16.9) | 22.7 | (22.4, 23.1) | 7.2 | (7.0, 7.4) | 34.8 | (34.4, 35.2) | 18.7 | (18.5, 19.1) |
| 2010 | 16.6 | (16.3, 16.9) | 20.3 | (20.0, 20.7) | 6.2 | (6.0, 6.4) | 36.3 | (35.9, 36.7) | 20.6 | (20.3, 20.9) |
| 2011 | 12.4 | (12.1, 12.7) | 22.9 | (22.6, 23.2) | 5.9 | (5.7, 6.1) | 36.7 | (36.3, 37.1) | 22.1 | (21.8, 22.4) |
| 2012 | 12.4 | (12.1, 12.7) | 21.8 | (21.4, 22.1) | 5.5 | (5.4, 5.8) | 36.8 | (36.4, 37.2) | 23.5 | (23.2, 23.9) |
| 2013 | 10.1 | (9.9, 10.4) | 22.1 | (21.8, 22.4) | 4.8 | (4.7, 5.0) | 38.1 | (37.7, 38.5) | 24.8 | (24.5, 25.2) |
| 2014 | 13.1 | (12.8, 13.5) | 17.1 | (16.8, 17.5) | 6.5 | (6.3, 6.7) | 37.5 | (37.1, 38.0) | 25.8 | (25.4, 26.2) |
| 2015 | 16.1 | (15.7, 16.4) | 13.4 | (13.1, 13.7) | 10.9 | (10.6, 11.2) | 33.2 | (32.7, 33.6) | 26.4 | (26.1, 26.8) |

Abbreviations: ART, antiretroviral therapy; CI, confidence interval

Table S6. Weighted prevalence of each level of the HIV care cascade and cumulative incidence of mortality by end of the calendar year, using an adherence level of 90%

| Calendar Year | Not retained Not ART adherent | | Retained Not ART adherent | | Not retained ART adherent | | Retained ART adherent | | Mortality Incidence | |
|---|---|---|---|---|---|---|---|---|---|---|
| | % | 95% CI | % | 95% CI | % | 95% CI | % | 95% CI | % | 95% CI |
| 2002 | 25.7 | (25.4, 26.1) | 46.6 | (46.2, 47.0) | 4.7 | (4.5, 4.9) | 18.7 | (18.4, 19.0) | 4.3 | (4.1, 4.4) |
| 2003 | 26.3 | (26.0, 26.7) | 41.7 | (41.3, 42.1) | 5.0 | (4.8, 5.2) | 20.3 | (20.0, 20.6) | 6.6 | (6.4, 6.8) |
| 2004 | 25.2 | (24.8, 25.5) | 38.7 | (38.3, 39.1) | 6.1 | (5.9, 6.3) | 21.0 | (20.7, 21.3) | 9.0 | (8.8, 9.2) |
| 2005 | 25.4 | (25.0, 25.7) | 36.3 | (35.9, 36.7) | 6.1 | (5.9, 6.3) | 20.9 | (20.7, 21.3) | 11.3 | (11.0, 11.5) |
| 2006 | 23.7 | (23.4, 24.1) | 32.0 | (31.6, 32.3) | 7.1 | (6.9, 7.3) | 23.8 | (23.5, 24.2) | 13.4 | (13.1, 13.7) |
| 2007 | 24.4 | (24.0, 24.7) | 28.9 | (28.6, 29.3) | 7.7 | (7.5, 7.9) | 23.8 | (23.5, 24.2) | 15.1 | (14.9, 15.4) |
| 2008 | 20.5 | (20.2, 20.8) | 29.4 | (29.0, 29.7) | 6.7 | (6.5, 6.9) | 26.5 | (26.1, 26.9) | 16.9 | (16.7, 17.3) |
| 2009 | 17.8 | (17.5, 18.1) | 28.7 | (28.4, 29.1) | 5.9 | (5.8, 6.1) | 28.8 | (28.4, 29.1) | 18.7 | (18.5, 19.1) |
| 2010 | 17.8 | (17.4, 18.1) | 26.7 | (26.3, 27.1) | 5.0 | (4.8, 5.2) | 30.0 | (29.6, 30.3) | 20.6 | (20.3, 20.9) |
| 2011 | 13.5 | (13.2, 13.8) | 29.5 | (29.1, 29.8) | 4.8 | (4.6, 4.9) | 30.1 | (29.8, 30.5) | 22.1 | (21.8, 22.4) |
| 2012 | 13.5 | (13.3, 13.8) | 28.8 | (28.5, 29.2) | 4.4 | (4.3, 4.6) | 29.7 | (29.3, 30.0) | 23.5 | (23.2, 23.9) |
| 2013 | 11.0 | (10.7, 11.3) | 28.7 | (28.3, 29.0) | 4.0 | (3.8, 4.1) | 31.5 | (31.1, 31.9) | 24.9 | (24.5, 25.2) |
| 2014 | 14.2 | (13.9, 14.6) | 23.3 | (22.9, 23.7) | 5.2 | (5.0, 5.5) | 31.5 | (31.1, 31.9) | 25.8 | (25.5, 26.2) |
| 2015 | 18.1 | (17.7, 18.4) | 18.9 | (18.6, 19.3) | 8.8 | (8.6, 9.1) | 27.7 | (27.4, 28.1) | 26.4 | (26.1, 26.8) |

Abbreviations: ART, antiretroviral therapy; CI, confidence interval

Table S7. Crude prevalence of each level of the HIV care cascade and cumulative incidence of mortality by end of the calendar, using an adherence level of 80%

| Calendar Year | Not retained Not ART adherent | | Retained Not ART adherent | | Not retained ART adherent | | Retained ART adherent | | Mortality Incidence | |
|---|---|---|---|---|---|---|---|---|---|---|
| | % | 95% CI | % | 95% CI | % | 95% CI | % | 95% CI | % | 95% CI |
| 2002 | 24.9 | (24.5, 25.2) | 42.9 | (42.5, 43.3) | 5.7 | (5.5, 5.9) | 22.4 | (22.0, 22.7) | 4.2 | (4.0, 4.3) |
| 2003 | 25.3 | (25.0, 25.7) | 37.7 | (37.3, 38.0) | 6.1 | (5.9, 6.3) | 24.5 | (24.2, 24.8) | 6.5 | (6.3, 6.6) |
| 2004 | 24.1 | (23.8, 24.5) | 34.8 | (34.4, 35.1) | 7.3 | (7.1, 7.5) | 25.0 | (24.7, 25.3) | 8.8 | (8.5, 9.0) |
| 2005 | 24.5 | (24.1, 24.8) | 32.2 | (31.9, 32.6) | 7.5 | (7.3, 7.7) | 24.8 | (24.5, 25.2) | 11.0 | (10.7, 11.2) |
| 2006 | 22.7 | (22.4, 23.0) | 27.4 | (27.1, 27.8) | 8.7 | (8.5, 8.9) | 28.2 | (27.8, 28.5) | 13.0 | (12.8, 13.3) |
| 2007 | 22.2 | (21.9, 22.6) | 25.1 | (24.7, 25.4) | 9.6 | (9.3, 9.8) | 28.4 | (28.0, 28.7) | 14.7 | (14.5, 15.0) |
| 2008 | 18.5 | (18.2, 18.8) | 24.9 | (24.6, 25.2) | 8.4 | (8.2, 8.7) | 31.7 | (31.3, 32.1) | 16.5 | (16.2, 16.8) |
| 2009 | 15.7 | (15.4, 16.0) | 24.1 | (23.8, 24.4) | 7.4 | (7.2, 7.6) | 34.5 | (34.2, 34.9) | 18.2 | (17.9, 18.5) |
| 2010 | 15.1 | (14.8, 15.3) | 22.7 | (22.4, 23.0) | 6.0 | (5.8, 6.2) | 36.1 | (35.8, 36.5) | 20.1 | (19.8, 20.4) |
| 2011 | 12.5 | (12.2, 12.8) | 25.3 | (25.0, 25.7) | 5.5 | (5.3, 5.7) | 35.0 | (34.7, 35.4) | 21.6 | (21.3, 22.0) |
| 2012 | 12.3 | (12.0, 12.5) | 23.8 | (23.5, 24.1) | 5.1 | (4.9, 5.3) | 35.8 | (35.4, 36.1) | 23.1 | (22.7, 23.4) |
| 2013 | 9.9 | (9.7, 10.1) | 24.2 | (23.9, 24.5) | 4.5 | (4.3, 4.6) | 36.9 | (36.6, 37.3) | 24.5 | (24.1, 24.8) |
| 2014 | 13.6 | (13.3, 14.0) | 18.5 | (18.2, 18.8) | 6.3 | (6.1, 6.5) | 36.1 | (35.7, 36.5) | 25.4 | (25.1, 25.8) |
| 2015 | 16.6 | (16.3, 17.0) | 14.8 | (14.5, 15.1) | 10.3 | (10.0, 10.6) | 32.2 | (31.8, 32.6) | 26.1 | (25.7, 26.4) |

Abbreviations: ART, antiretroviral therapy; CI, confidence interval

Table S8. Crude prevalence of each level of the HIV care cascade and cumulative incidence of mortality by end of the calendar year, using an adherence level of 0%

| Calendar Year | Not retained Not ART adherent | | Retained Not ART adherent | | Not retained ART adherent | | Retained ART adherent | | Mortality Incidence | |
|---|---|---|---|---|---|---|---|---|---|---|
| | % | 95% CI | % | 95% CI | % | 95% CI | % | 95% CI | % | 95% CI |
| 2002 | 22.0 | (21.6, 22.3) | 34.1 | (33.7, 34.5) | 8.5 | (8.3, 8.7) | 31.2 | (30.9, 31.6) | 4.2 | (4.0, 4.3) |
| 2003 | 22.6 | (22.3, 23.0) | 29.5 | (29.2, 29.9) | 8.7 | (8.4, 8.9) | 32.7 | (32.4, 33.1) | 6.4 | (6.2, 6.6) |
| 2004 | 21.7 | (21.3, 22.0) | 26.9 | (26.6, 27.3) | 9.8 | (9.6, 10.1) | 32.9 | (32.5, 33.2) | 8.7 | (8.5, 9.0) |
| 2005 | 21.9 | (21.6, 22.3) | 24.7 | (24.3, 25.0) | 10.1 | (9.8, 10.3) | 32.4 | (32.0, 32.7) | 10.9 | (10.7, 11.2) |
| 2006 | 20.0 | (19.7, 20.3) | 19.5 | (19.2, 19.8) | 11.5 | (11.2, 11.7) | 36.0 | (35.6, 36.3) | 13.0 | (12.8, 13.3) |
| 2007 | 19.6 | (19.3, 20.0) | 17.5 | (17.2, 17.8) | 12.3 | (12.0, 12.6) | 35.9 | (35.5, 36.2) | 14.7 | (14.4, 15.0) |
| 2008 | 16.6 | (16.2, 16.9) | 17.3 | (17.0, 17.6) | 10.5 | (10.3, 10.8) | 39.2 | (38.8, 39.6) | 16.5 | (16.2, 16.7) |
| 2009 | 14.0 | (13.7, 14.3) | 16.2 | (15.9, 16.4) | 9.3 | (9.0, 9.5) | 42.3 | (41.9, 42.7) | 18.2 | (17.9, 18.5) |
| 2010 | 13.4 | (13.1, 13.7) | 14.3 | (14.0, 14.5) | 7.8 | (7.6, 8.0) | 44.5 | (44.1, 44.9) | 20.1 | (19.8, 20.4) |
| 2011 | 11.0 | (10.7, 11.2) | 15.9 | (15.6, 16.1) | 7.1 | (6.9, 7.3) | 44.4 | (44.0, 44.8) | 21.6 | (21.3, 21.9) |
| 2012 | 10.3 | (10.1, 10.6) | 13.6 | (13.4, 13.9) | 7.0 | (6.8, 7.2) | 46.0 | (45.6, 46.4) | 23.0 | (22.7, 23.4) |
| 2013 | 8.4 | (8.2, 8.6) | 14.8 | (14.6, 15.1) | 5.8 | (5.6, 6.0) | 46.5 | (46.2, 46.9) | 24.4 | (24.1, 24.8) |
| 2014 | 12.0 | (11.7, 12.3) | 10.7 | (10.4, 10.9) | 8.1 | (7.8, 8.3) | 43.9 | (43.4, 44.3) | 25.4 | (25.1, 25.8) |
| 2015 | 14.1 | (13.8, 14.4) | 8.1 | (7.9, 8.3) | 13.1 | (12.8, 13.4) | 38.7 | (38.3, 39.1) | 26.0 | (25.7, 26.4) |

Abbreviations: ART, antiretroviral therapy; CI, confidence interval

Table S9. Crude prevalence of each level of the HIV care cascade and cumulative incidence of mortality by end of the calendar year, using an adherence level of 70%

| Calendar Year | Not retained Not ART adherent | | Retained Not ART adherent | | Not retained ART adherent | | Retained ART adherent | | Mortality Incidence | |
|---|---|---|---|---|---|---|---|---|---|---|
| | % | 95% CI | % | 95% CI | % | 95% CI | % | 95% CI | % | 95% CI |
| 2002 | 24.4 | (24.0, 24.7) | 41.1 | (40.7, 41.5) | 6.2 | (6.0, 6.4) | 24.2 | (23.9, 24.5) | 4.2 | (4.0, 4.3) |
| 2003 | 24.8 | (24.5, 25.2) | 35.9 | (35.5, 36.3) | 6.5 | (6.3, 6.7) | 26.3 | (25.9, 26.6) | 6.4 | (6.3, 6.6) |
| 2004 | 23.5 | (23.2, 23.9) | 32.8 | (32.4, 33.1) | 7.9 | (7.7, 8.1) | 27.0 | (26.7, 27.4) | 8.8 | (8.5, 9.0) |
| 2005 | 23.8 | (23.5, 24.2) | 30.3 | (29.9, 30.6) | 8.1 | (7.9, 8.3) | 26.8 | (26.5, 27.2) | 11.0 | (10.7, 11.2) |
| 2006 | 22.0 | (21.6, 22.3) | 25.2 | (24.9, 25.5) | 9.4 | (9.2, 9.7) | 30.4 | (30.1, 30.8) | 13.0 | (12.8, 13.3) |
| 2007 | 21.5 | (21.1, 21.8) | 23.0 | (22.6, 23.3) | 10.4 | (10.1, 10.6) | 30.5 | (30.1, 30.8) | 14.7 | (14.4, 15.0) |
| 2008 | 18.1 | (17.7, 18.4) | 23.2 | (22.8, 23.5) | 8.9 | (8.6, 9.1) | 33.4 | (33.1, 33.8) | 16.5 | (16.2, 16.8) |
| 2009 | 15.3 | (15.0, 15.6) | 22.2 | (21.9, 22.5) | 7.9 | (7.6, 8.1) | 36.4 | (36.0, 36.8) | 18.2 | (17.9, 18.5) |
| 2010 | 14.6 | (14.3, 14.9) | 20.3 | (20.0, 20.6) | 6.5 | (6.3, 6.7) | 38.5 | (38.1, 38.9) | 20.1 | (19.8, 20.4) |
| 2011 | 12.1 | (11.8, 12.3) | 23.0 | (22.7, 23.3) | 5.9 | (5.7, 6.1) | 37.4 | (37.0, 37.7) | 21.6 | (21.3, 21.9) |
| 2012 | 11.8 | (11.6, 12.1) | 21.4 | (21.1, 21.7) | 5.5 | (5.3, 5.7) | 38.2 | (37.9, 38.6) | 23.1 | (22.7, 23.4) |
| 2013 | 9.6 | (9.4, 9.8) | 22.0 | (21.8, 22.3) | 4.8 | (4.6, 4.9) | 39.1 | (38.8, 39.5) | 24.4 | (24.1, 24.8) |
| 2014 | 13.2 | (12.9, 13.5) | 16.6 | (16.3, 16.9) | 6.8 | (6.5, 7.0) | 38.0 | (37.6, 38.4) | 25.4 | (25.1, 25.8) |
| 2015 | 16.1 | (15.7, 16.4) | 13.3 | (13.1, 13.6) | 10.9 | (10.6, 11.2) | 33.6 | (33.2, 34.0) | 26.1 | (25.7, 26.4) |

Abbreviations: ART, antiretroviral therapy; CI, confidence interval

Table S10. Crude prevalence of each level of the HIV care cascade and cumulative incidence of mortality by end of the calendar, using an adherence level of 90%

| Calendar Year | Not retained Not ART adherent | | Retained Not ART adherent | | Not retained ART adherent | | Retained ART adherent | | Mortality Incidence | |
|---|---|---|---|---|---|---|---|---|---|---|
| | % | 95% CI | % | 95% CI | % | 95% CI | % | 95% CI | % | 95% CI |
| 2002 | 25.8 | (25.4, 26.1) | 46.3 | (45.9, 46.7) | 4.8 | (4.6, 5.0) | 19.0 | (18.7, 19.3) | 4.2 | (4.0, 4.3) |
| 2003 | 26.3 | (25.9, 26.7) | 41.3 | (40.9, 41.7) | 5.1 | (4.9, 5.3) | 20.8 | (20.5, 21.1) | 6.5 | (6.3, 6.7) |
| 2004 | 25.1 | (24.8, 25.5) | 38.2 | (37.8, 38.6) | 6.3 | (6.1, 6.5) | 21.6 | (21.3, 21.9) | 8.8 | (8.5, 9.0) |
| 2005 | 25.5 | (25.2, 25.9) | 35.6 | (35.2, 36.0) | 6.4 | (6.2, 6.6) | 21.5 | (21.2, 21.8) | 11.0 | (10.7, 11.2) |
| 2006 | 23.8 | (23.5, 24.2) | 31.0 | (30.7, 31.4) | 7.5 | (7.3, 7.7) | 24.6 | (24.2, 24.9) | 13.0 | (12.8, 13.3) |
| 2007 | 23.4 | (23.1, 23.7) | 28.8 | (28.4, 29.1) | 8.4 | (8.2, 8.6) | 24.7 | (24.4, 25.0) | 14.7 | (14.5, 15.0) |
| 2008 | 19.5 | (19.2, 19.9) | 28.9 | (28.5, 29.2) | 7.4 | (7.2, 7.6) | 27.7 | (27.4, 28.1) | 16.5 | (16.2, 16.8) |
| 2009 | 16.6 | (16.3, 16.9) | 28.3 | (28.0, 28.7) | 6.5 | (6.3, 6.7) | 30.3 | (29.9, 30.7) | 18.2 | (17.9, 18.5) |
| 2010 | 15.8 | (15.5, 16.1) | 26.9 | (26.5, 27.2) | 5.3 | (5.1, 5.4) | 32.0 | (31.6, 32.3) | 20.1 | (19.8, 20.4) |
| 2011 | 13.2 | (12.9, 13.4) | 29.5 | (29.2, 29.8) | 4.8 | (4.7, 5.0) | 30.9 | (30.5, 31.2) | 21.6 | (21.3, 22.0) |
| 2012 | 13.0 | (12.7, 13.2) | 28.5 | (28.1, 28.8) | 4.4 | (4.2, 4.6) | 31.1 | (30.8, 31.5) | 23.1 | (22.7, 23.4) |
| 2013 | 10.4 | (10.2, 10.7) | 28.6 | (28.3, 29.0) | 4.0 | (3.8, 4.1) | 32.5 | (32.1, 32.8) | 24.5 | (24.1, 24.8) |
| 2014 | 14.4 | (14.1, 14.7) | 22.8 | (22.5, 23.2) | 5.5 | (5.3, 5.7) | 31.8 | (31.4, 32.2) | 25.5 | (25.1, 25.8) |
| 2015 | 18.0 | (17.7, 18.4) | 18.9 | (18.6, 19.2) | 8.8 | (8.6, 9.1) | 28.2 | (27.8, 28.5) | 26.1 | (25.7, 26.4) |

Abbreviations: ART, antiretroviral therapy; CI, confidence interval

Table S11. Weighted prevalence of each level of the HIV care cascade and cumulative incidence of mortality by end of the calendar year, using an adherence level of 80% and excluding California

| Calendar Year | Not retained Not ART adherent | | Retained Not ART adherent | | Not retained ART adherent | | Retained ART adherent | | Mortality Incidence | |
|---|---|---|---|---|---|---|---|---|---|---|
| | % | 95% CI | % | 95% CI | % | 95% CI | % | 95% CI | % | 95% CI |
| 2002 | 25.6 | (25.3, 26.0) | 42.7 | (42.3, 43.1) | 5.4 | (5.2, 5.6) | 21.5 | (21.1, 21.8) | 4.8 | (4.6, 5.0) |
| 2003 | 26.0 | (25.7, 26.4) | 37.8 | (37.4, 38.1) | 5.5 | (5.3, 5.7) | 23.3 | (22.9, 23.6) | 7.5 | (7.3, 7.7) |
| 2004 | 24.6 | (24.3, 24.9) | 35.0 | (34.6, 35.4) | 6.7 | (6.5, 6.9) | 23.5 | (23.1, 23.9) | 10.2 | (9.9, 10.5) |
| 2005 | 25.1 | (24.7, 25.4) | 32.4 | (32.0, 32.7) | 7.1 | (6.9, 7.3) | 22.7 | (22.4, 23.1) | 12.7 | (12.5, 13.0) |
| 2006 | 23.3 | (23.0, 23.6) | 27.9 | (27.6, 28.3) | 8.1 | (7.9, 8.3) | 25.5 | (25.2, 25.8) | 15.2 | (14.9, 15.5) |
| 2007 | 24.1 | (23.8, 24.4) | 24.4 | (24.1, 24.8) | 8.8 | (8.5, 9.0) | 25.6 | (25.2, 25.9) | 17.1 | (16.8, 17.5) |
| 2008 | 20.0 | (19.7, 20.4) | 25.0 | (24.6, 25.3) | 7.5 | (7.3, 7.7) | 28.4 | (28.0, 28.7) | 19.2 | (18.8, 19.5) |
| 2009 | 17.2 | (16.9, 17.6) | 24.2 | (23.9, 24.6) | 6.4 | (6.2, 6.6) | 30.9 | (30.6, 31.3) | 21.2 | (20.9, 21.6) |
| 2010 | 17.1 | (16.8, 17.5) | 22.4 | (22.0, 22.7) | 4.8 | (4.6, 5.0) | 32.4 | (32.0, 32.8) | 23.3 | (22.9, 23.7) |
| 2011 | 12.5 | (12.2, 12.8) | 25.6 | (25.2, 26.0) | 4.5 | (4.3, 4.7) | 32.5 | (32.0, 32.9) | 25.0 | (24.6, 25.3) |
| 2012 | 12.3 | (12.0, 12.6) | 24.4 | (24.1, 24.8) | 4.2 | (4.0, 4.3) | 32.5 | (32.1, 32.9) | 26.5 | (26.2, 26.9) |
| 2013 | 9.7 | (9.5, 10.0) | 24.5 | (24.2, 24.8) | 3.6 | (3.4, 3.7) | 34.2 | (33.8, 34.5) | 28.0 | (27.7, 28.4) |
| 2014 | 14.0 | (13.6, 14.4) | 18.7 | (18.4, 19.1) | 5.5 | (5.2, 5.7) | 32.6 | (32.1, 33.1) | 29.2 | (28.8, 29.6) |
| 2015 | 18.5 | (18.1, 19.0) | 14.1 | (13.8, 14.5) | 9.9 | (9.6, 10.3) | 27.3 | (26.9, 27.8) | 30.1 | (29.7, 30.5) |

Abbreviations: ART, antiretroviral therapy; CI, confidence interval

Table S12. Weighted prevalence of each level of the HIV care cascade and cumulative incidence of mortality by end of the calendar year, using an adherence level of 80% and using September 30, 2015 as the administrative censoring date for all states

| Calendar Year | Not retained Not ART adherent | | Retained Not ART adherent | | Not retained ART adherent | | Retained ART adherent | | Mortality Incidence | |
|---|---|---|---|---|---|---|---|---|---|---|
| | % | 95% CI | % | 95% CI | % | 95% CI | % | 95% CI | % | 95% CI |
| 2002 | 25.0 | (24.6, 25.3) | 43.2 | (42.8, 43.5) | 5.6 | (5.4, 5.8) | 22.0 | (21.7, 22.3) | 4.3 | (4.1, 4.4) |
| 2003 | 25.7 | (25.4, 26.1) | 37.9 | (37.6, 38.2) | 6.0 | (5.8, 6.1) | 23.7 | (23.4, 24.1) | 6.6 | (6.5, 6.8) |
| 2004 | 24.6 | (24.2, 24.9) | 35.0 | (34.6, 35.3) | 7.3 | (7.1, 7.5) | 24.2 | (23.8, 24.5) | 9.0 | (8.8, 9.3) |
| 2005 | 24.7 | (24.4, 25.1) | 32.7 | (32.3, 33.1) | 7.3 | (7.1, 7.5) | 24.0 | (23.7, 24.4) | 11.3 | (11.1, 11.6) |
| 2006 | 22.9 | (22.6, 23.2) | 28.2 | (27.9, 28.6) | 8.2 | (8.1, 8.4) | 27.2 | (26.9, 27.5) | 13.5 | (13.2, 13.7) |
| 2007 | 23.4 | (23.1, 23.8) | 25.1 | (24.7, 25.4) | 8.9 | (8.7, 9.1) | 27.4 | (27.0, 27.7) | 15.2 | (14.9, 15.5) |
| 2008 | 19.6 | (19.2, 20.0) | 25.4 | (25.1, 25.8) | 7.7 | (7.5, 7.9) | 30.2 | (29.8, 30.6) | 17.0 | (16.7, 17.4) |
| 2009 | 17.2 | (16.8, 17.5) | 24.4 | (24.1, 24.8) | 6.9 | (6.7, 7.0) | 32.7 | (32.3, 33.0) | 18.8 | (18.5, 19.2) |
| 2010 | 17.3 | (17.0, 17.6) | 22.4 | (22.1, 22.7) | 6.0 | (5.8, 6.2) | 33.7 | (33.3, 34.0) | 20.7 | (20.4, 21.0) |
| 2011 | 13.3 | (13.0, 13.6) | 24.9 | (24.6, 25.3) | 5.7 | (5.5, 6.0) | 33.9 | (33.5, 34.2) | 22.2 | (21.9, 22.5) |
| 2012 | 13.2 | (12.9, 13.5) | 23.9 | (23.5, 24.2) | 5.4 | (5.2, 5.6) | 33.9 | (33.5, 34.2) | 23.6 | (23.3, 23.9) |
| 2013 | 10.7 | (10.4, 11.0) | 25.2 | (24.9, 25.5) | 4.6 | (4.5, 4.8) | 34.5 | (34.1, 34.8) | 25.0 | (24.7, 25.3) |
| 2014 | 11.7 | (11.4, 11.9) | 27.4 | (27.1, 27.7) | 4.5 | (4.4, 4.7) | 30.5 | (30.2, 30.8) | 25.9 | (25.6, 26.3) |
| 2015 | 14.1 | (13.9, 14.3) | 27.6 | (27.4, 27.9) | 6.8 | (6.6, 6.9) | 25.0 | (24.8, 25.3) | 26.5 | (26.1, 26.8) |

Abbreviations: ART, antiretroviral therapy; CI, confidence interval
